# A wireless magnetic implant system for continuous neuromuscular sensing

**DOI:** 10.1101/2025.11.17.25340274

**Authors:** Christopher C. Shallal, Cameron R. Taylor, Seong Ho Yeon, Richard J. Casler, Dara Oseyemi, Guillermo Herrera-Arcos, Junqing Qiao, Tony Shu, Jay Xu, Daniel Levine, Ana Rajcevic, Sean Boerhout, Aimee Liu, Ellen G. Clarrissimeaux, Joseph A. Paradiso, Matthew J. Carty, Hugh M. Herr

## Abstract

Precise measurement of neuromuscular mechanics yields an intuitive control signal for producing synchronous movement with wearable robotics. Practically obtaining such measurements remains challenging as current muscle-sensing technologies excessively compromise between signal fidelity, system complexity, and invasiveness. Here we present a skin-mounted, magnetometer array platform that wirelessly tracks passive 3 mm diameter magnetic beads implanted within human muscle tissue for continuous neuromuscular sensing. The system employs customized high-density sensing electronics with an information-theoretic architecture to achieve sub-millimeter resolution of real-time muscle dynamics at tracking depths of up to 6 cm within the body. We deploy the platform in a first-in-human clinical study to track a constellation of permanently implanted magnets to enable multiple degree-of-freedom neuroprosthetic control. We demonstrate that the wireless muscle state estimation can outperform standard surface and implanted electromyography interfaces to achieve more accurate and responsive neuroprosthetic movement. Further, we successfully supplant electromyography altogether by extending the platform to detect muscle activation through magnetic induction alone.

**One-Sentence Summary:** A skin-mounted magnetometer sensing array can wirelessly track 3 mm diameter permanent magnetic implants in humans to provide precise neuromuscular information for improved neuroprosthetic control compared to electromyography.

## Main

Scientists, technologists, and clinicians have long sought neural interfaces that enable reproducible and repeatable signaling between the peripheral nervous system and wearable robotics designed to restore lost biological capabilities^1,2^. Electromyography (EMG), where electrodes record muscle electrical activity to infer user intent, has been the predominant approach for neuroprosthetic control^3,4^. Despite considerable research focused on enabling more versatile, direct neural control of movements, widespread clinical adoption has remained elusive due to major limitations^5,6^. EMG monitors electrical rather than mechanical signals, requiring complex models and continual calibration to infer muscle state or force^7–9^. Additionally, current surface EMG (sEMG) sensing platforms suffer from electrode–skin liftoff, movement artifacts and changes in skin impedance that limit performance outside controlled laboratory environments^10,11^. Implanted EMG (iEMG) mitigates these shortcomings but demands highly invasive surgery with internal wiring and associated medical risks^12,13^. Current research that focuses on neuroprosthetic interfacing involving new surgical techniques, powered bionic prostheses and neural control still lacks a viable and efficacious sensing interface^14–16^.

Sensing directly in the mechanical domain offers more interpretable and physiologically-aligned access to tissue dynamics^17,18^. Sonomicrometry and fluoromicrometry can measure muscle dynamics^19–21^, but sonomicrometry requires highly invasive tethered implants, fluoromicrometry involves radiation exposure, and neither technique is fully portable. Ultrasound is actively being explored as a real-time sensing interface, with work also ongoing to improve transducer portability^22–25^. B-mode ultrasound tracking requires substantial signal processing, bulky probes and is insufficiently accurate for reliable neuroprosthetic control^26^. A-mode approaches, while showing promise for neuroprosthetic applications, require a large number of transducer channels for optimal acquisition and rely on trained models for classification rather than direct kinematic measurements^27^.

Magnetomicrometry and other magnetic-based measurement techniques that spatially track an implanted magnet with an external sensor have been developed for muscle length sensing applications in both animals^28–30^ and humans^31,32^. However, constraints such as low signal-to-noise ratios and shallow tracking depths (<3 cm) prevent placement in deeper tissue and limit the number of implantable magnets, restricting accessible muscle signals and neuroprosthetic functionality. Additionally, these interfaces required machine learning compensation or only tracked magnetic field changes rather than computed magnet positions, precluding accurate interpretation of full muscle state and demonstrating no significant performance improvements over standard EMG. Furthermore, previous work in humans has required larger magnets and been followed quickly by explantation, limiting clinical implementation to case studies. Current systems have not achieved the simultaneous requirements for high-fidelity neuroprosthetic control: smaller implants, low latency, precise localization, deep tissue tracking, and multi-magnet capability (Supplementary Table 1).

In this Article, we present a wearable, wireless muscle state estimator (MuSE) that enables simultaneous tracking of multiple 3 mm diameter magnetic beads implanted in muscle with sub-millimeter resolution at depths of up to 6 cm (Fig. 1a,b). To capture full muscle state in humans, we design custom electronics consisting of a high-density magnetic field sensor architecture driven by a field-programmable gate array (FPGA) for high-bandwidth, parallel measurements. We formulate a computationally efficient localization algorithm based on information theory that exploits the multiplexed sensing to decode precise magnet positions from measured field data. Continuous in vivo monitoring of muscle tissue dynamics is estimated from the magnet movement (Fig. 1c). To demonstrate the MuSE system’s capabilities, we conduct a human clinical study on three participants with transtibial amputation. Up to six magnets are implanted in four residual limb muscles to enable direct neuroprosthetic control. These same four muscles are also instrumented with both invasive and skin-mounted EMG electrodes. Through this augmented neuroprosthetic interface, the MuSE generates more interpretable, mechanically representative, and accurate control signals compared to both the surface and invasive EMG interfaces. During neuroprosthetic tasks, the MuSE interface sustains stable, complex, and multiple degree-of-freedom (DOF) control that outperforms the EMG interfaces. We further leverage principles of magnetic induction to enable contactless muscle activation sensing through a compact auxiliary coil, demonstrating the potential for comprehensive muscle state monitoring.

**Fig. 1.**
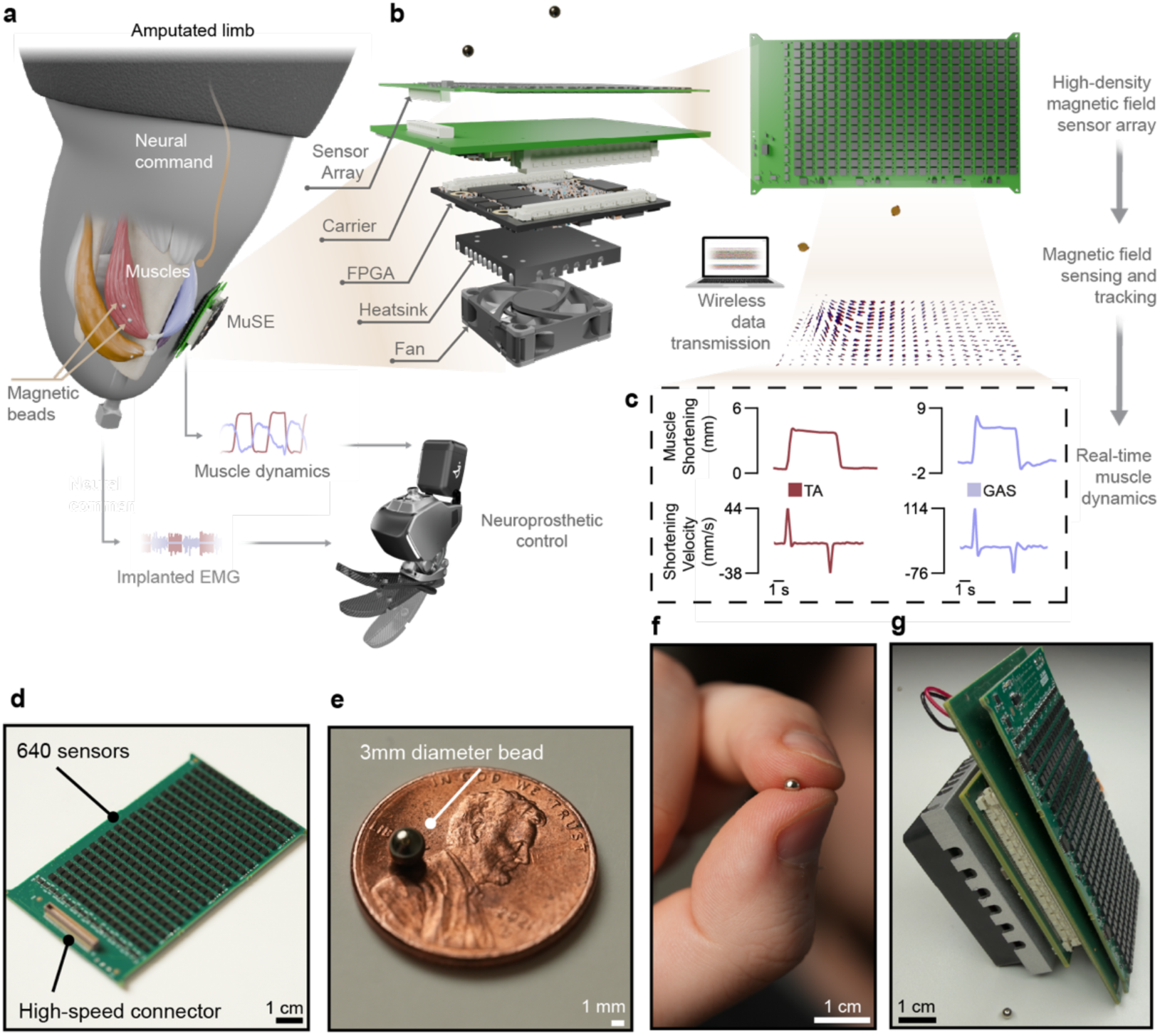
Wireless MuSE overview. **a,** Schematic and working principle of MuSE where implanted magnets are placed in the residual muscles of an amputated limb to track muscle dynamics. The MuSE wirelessly provides measurements in real-time which encompass the muscle state. These signals are used to infer joint dynamics for controlling either a robotic or virtual neuroprosthetic limb and are compared to standard or implanted muscle sensing interfaces such as EMG. **b,** Illustration of the MuSE system where embedded electronics consist of a 640 magnetic field sensor board linked to an FPGA computing module through a custom carrier PCB that samples all 640 sensors at 1 kHz to track the magnets. The magnetic field information is then aggregated through a tracking algorithm that solves the inverse dipole problem to map from the measured fields to the magnet states. **c,** Measurement of the relative distance between two magnets within the same muscle belly enabled extraction of muscle tissue length and velocity (for example, in the gastrocnemius (GAS) and tibialis anterior (TA) muscles), and these muscle dynamic signals can be sent to a bionic prosthesis as neuromuscular control inputs. **d,** Photograph of high-density sensor board that tracks the beads from outside the body, which is no larger than a credit card. **e-f,** Images of the small size (3 mm diameter) implanted magnetic beads with penny for scale. **g,** Depiction of entire embedded electronic system with all components and magnetic bead for scale.

### Wireless MuSE system design

The MuSE system integrates four core components: (i) an external high-density magnetometer array that measures the magnetic fields generated by magnetic implants, (ii) passive magnetic implants placed within muscle tissue, (iii) embedded high-bandwidth state estimation that outputs magnet movement using physics-based models and algorithms, and (iv) real-time computation that derives muscle-tendon dynamics from magnet states for neuroprosthetic control (Fig. 1a-c). The external sensor array is a custom printed circuit board (PCB) comprising 640 three-axis magnetometers (LIS3MDL, STMicroelectronics) arranged in a rectangular 53 mm × 86 mm grid, providing high spatial resolution for tracking implanted magnets and allowing a small, dense footprint for the embedded system (Fig. 1d). The PCB features sensors on both top and bottom surfaces, with each surface containing a 20 × 16 grid (320 sensors per surface, 640 total). A main challenge in high-density and high-throughput sensing is careful management of data streams to ensure synchronization and signal quality. At a target 10 MHz clock frequency for the sensor communication protocol, standard logic buffers cannot provide sufficient current to maintain acceptable signal integrity. Custom electronics allow the sensors to stream data through a multiplexed communication architecture with dedicated clock distribution and row-sequential addressing to manage the high-density array. (Methods and Supplementary Fig. 1-2). The number of sensors was chosen to maximize sensor density given the required spatial dimensions, and sensors were placed on both sides such that the footprint would remain small while doubling the sensor count. Sensors were spaced 2.7 mm apart in the 16-row direction and 3.6 mm apart in the 20-row direction. Each sensor captures real-time three-dimensional magnetic field vectors generated by the implanted beads, with the dense multiplexed array measuring the spatial variation of the field. The magnetometers are calibrated using hard and soft iron calibration techniques to remove local magnetic biases and sensor-specific offsets.

The implanted markers are spherical neodymium-based magnetic beads (Halifax Biomedical), approximately 3 mm in diameter, with an N48SH rating providing residual flux density between 1.36 T and 1.42 T (Fig. 1e,f). These passive implants require no power or electronics, allowing for permanent implantation without concerns for battery life or device failure. To ensure long-term biocompatibility and stability, each bead was coated with a tri-layer system: a 10 μm Ni-Cu-Ni base layer for corrosion resistance, 5 μm of 99.9% pure gold for biocompatibility, and a 21 μm Parylene C outer coating for tissue integration. This size balances minimal tissue disruption with sufficient magnetic field strength for reliable tracking, as smaller beads would compromise signal quality while larger magnets increase surgical invasiveness. The MuSE system’s 3 mm diameter spherical magnetic beads are substantially smaller than magnets used in other work, promoting clinical translatability. (Supplementary Table 1).

The system incorporates an FPGA computing module (Mercury XU5, Enclustra) that controls sensor sampling at 1 kHz and processes the multiplexed magnetic field data in real-time. A custom carrier PCB connects the FPGA to the sensor array and provides communication interfaces for wireless or Ethernet data streaming to a neuroprosthetic interface (Fig. 1g). The MuSE system is powered by an external 12 V supply and can be battery-operated, with the sensor array and FPGA consuming approximately 0.9 A during continuous operation, while thermal management through heatsinks and small active cooling fans prevents magnetometer drift during extended use. The MuSE system’s modular architecture enables dynamic adjustment of both sampling rate and active sensor count to optimize power consumption, which can be reduced by half, based on magnet depth and required tracking precision.

The MuSE design enables scalable deployment of multiple sensor arrays for expanded spatial coverage or increased tracking precision (Supplementary Fig. 3-4 and Supplementary Note 1). This arrangement can either synchronize magnetic field measurements across all deployed boards for enhanced signal processing or independently track multiple moving tissues simultaneously. To coordinate multiple arrays, we augmented the traditional magnetometer calibration process with orientation and position calibration to determine the relative pose of each array (Supplementary Fig. 5). Magnetometer orientations were determined by applying a pure-rotation point cloud registration using an optimization algorithm after magnetometer calibration (Supplementary Fig. 6-7). We then independently tracked a calibration magnet from each array and determined array positions via pure-translation point cloud registration (Supplementary Fig. 8-10). Each board executes its own independent tracking algorithm, operating either within its local reference frame or within a global coordinate system following pose calibration, allowing multiple MuSE systems to function seamlessly together as a unified, scalable sensing platform.

### Tracking characterization and spatial resolution

The MuSE system’s high-density sensor array and high-bandwidth FPGA architecture provide the electronic hardware foundation for precise multi-magnet tracking, but utilizing this capability requires a complementary algorithmic framework that can efficiently process the multiplexed sensor data in real-time. Tracking multiple magnets implanted at varying depths across different muscles presents a significant computational challenge. Traditional approaches using optimization methods such as the Levenberg-Marquardt (LM) algorithm solve the inverse dipole problem independently at each time step, which injects sensor noise directly into position estimates and becomes computationally expensive when tracking multiple magnets simultaneously^33,34^. To address these limitations, we developed an information filter (INFO) framework that propagates uncertainty through precision matrices, explicitly models sensor noise characteristics, and defines temporal correlations in magnet positions through second-order dynamics modeling (Fig. 2a, Supplementary Fig. 11 and Supplementary Note 2). This approach leverages the fact that muscle tissue dynamics are correlated across time, allowing sequential measurements to refine position estimates rather than treating each timepoint independently. The information-theoretic formulation scales efficiently with sensor count and enables principled uncertainty quantification across varying sensor and magnet configurations.

**Fig. 2.**
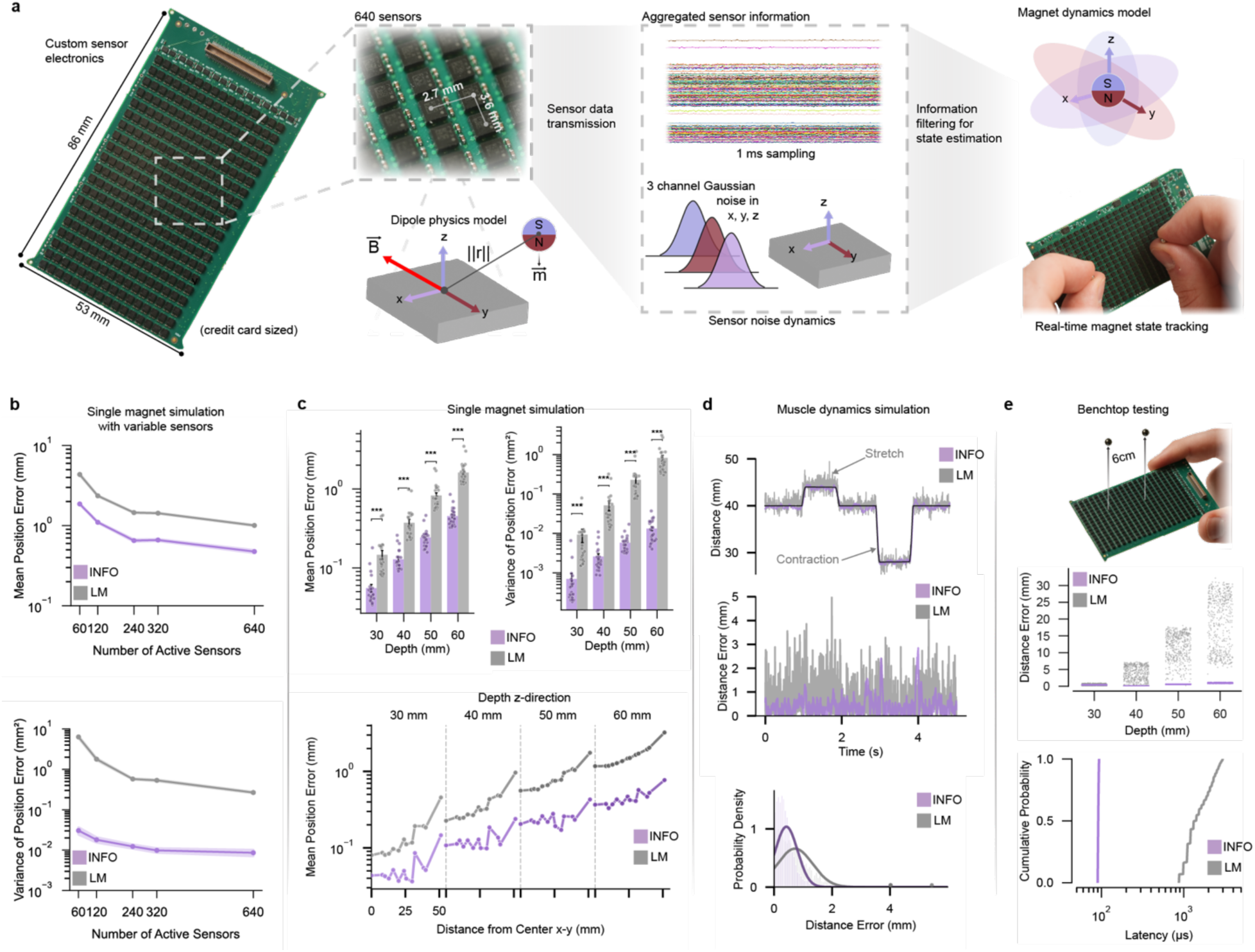
MuSE architecture with information theory. **a,** Information filter-based tracking algorithm (INFO) that takes incoming aggregated sensor information and uses a dipole physics model to solve the inverse problem, computing the position and orientation of the magnetic bead. The tracking algorithm incorporated both sensor and magnet dynamics to reduce noise and variability in the estimation output. The INFO was compared to the standard Levenberg-Marquardt (LM) optimization approach. **b,** Simulated test case to validate the impact of sensor number on estimating magnet state (n = 10 trials per sensor number). Increasing sensor number reduced mean position error and error variance for both LM and INFO algorithms. **c,** Simulated test case involving the maximum number of sensors and a single static magnet at varying x-, y-, and z-positions. Comparison of estimation error and error variance between the two algorithms demonstrated lower error and variance for INFO compared to LM (***P < 0.001 for each depth). **d,** Muscle dynamics simulation of both muscle stretch and contraction to evaluate each algorithm’s capability to track realistic, fast dynamics at 5 cm depth. Distance between the two-magnet trajectory reference is overlaid with INFO and LM algorithm outputs. Error across trajectory time and Gaussian distribution of error are also shown. **e,** Benchtop results, with real-time tracking algorithm implementations on the embedded system tracking the distance between two static magnets at varying matched z-depths. Strip plots demonstrate lower distance error with increasing depth for both LM and INFO algorithms. INFO has lower algorithm latency compared to LM. Statistical tests to determine significance was Mann Whitney U-test (Wilcoxon rank-sum). Bars represent mean and SEM. Shaded regions indicate SEM.

We validated the tracking framework through both simulation studies and benchtop experiments. A virtual environment reproduced sensor measurements by generating expected magnetic fields from known magnet positions, with simulations injecting 2.5 μT mean sensor noise while tracking magnetic beads with 1.39 T residual flux density at 1 kHz sampling rate— matching the physical system specifications. To comprehensively evaluate performance, two sets of simulation experiments were conducted: first, a single magnet at a fixed position was tracked while systematically varying active sensor count from 60 to 640 magnetometers to evaluate how sensor density affects tracking performance; second, using the full 640 sensor array, a single magnet was tracked across 44 spatial configurations spanning four depths (30-60 mm) with 11 different positions per depth to assess accuracy and precision across the sensing environment. Simulation studies that varied sensor density demonstrated that both INFO and LM algorithms improved with increasing sensor density, showing reduced position error as sensor count increased (Fig. 2b). The INFO consistently outperformed LM across all configurations, achieving lower mean position errors and reduced variance. The relationship between precision and sensor count follows a logarithmic scaling law 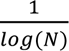 with *N* number of sensors, indicating that sensor measurements exhibit spatial correlation due to the dipole field structure and that each sensor contributes additively to the information matrix. With the full number of sensors and at depths of 30–60 mm across different magnet positions, INFO maintained sub-millimeter tracking accuracy while LM tracking errors exceeded 2 mm (Fig. 2c and Supplementary Fig. 12). The performance gap widens with depth and subsequent decreasing magnetic field strength, with INFO demonstrating significantly lower errors and variance at all depths tested (***P < 0.001) (Fig. 2c and Supplementary Fig. 13). Dynamic muscle simulation confirmed INFO’s superior tracking of physiological movements, achieving substantially lower mean tracking error (0.42 mm vs 0.72 mm) and improved precision (0.38 mm vs 0.60 mm standard deviation) compared to LM (Fig. 2d). These results demonstrate that the INFO effectively exploits the MuSE system’s high-density sensor array and 1 kHz sampling rate.

Benchtop validation using two physical magnets positioned 40 mm apart at depths ranging from 30-60 mm confirmed the computational advantages observed in simulation. INFO processing required 98 μs per update at the 99th percentile compared to 5530 μs for LM, representing more than a 50-fold speedup while simultaneously achieving lower estimation errors across all tested depths (Fig. 2e). The LM method at depths beyond 30 mm began to have millimeter-scale errors, while INFO maintained sub-millimeter errors (Supplementary Video 1). Given that physiologically relevant muscle deflections occur at the millimeter scale, the errors and high variance inherent in LM rendered such methods unsuitable for accurate muscle state estimation. This computational efficiency gain indicated that an improvement in algorithm architecture need not compromise real-time tracking ability. The combination of temporal dynamics modeling and efficient information matrix computations allowed INFO to successfully track magnets up to 6 cm deep with sub-millimeter accuracy, approximately doubling the tracking depth of previous magnetic sensing systems while maintaining the precision necessary for robust muscle state estimation (Supplementary Video 2).

### Clinical validation of MuSE system with existing neuromuscular interfaces

Having characterized the MuSE system’s performance in tracking magnets, we determined its clinical utility in providing continuous muscle dynamics and compared the signals to currently used sensing interfaces. We recruited three participants with transtibial amputations who had undergone a bionic reconstruction. Each participant received an osseointegrated implant system integrating both implanted electrodes and magnetic beads (Fig. 3a). The beads were implanted into residual muscles—two magnets per ankle muscle and one per subtalar muscle due to spatial constraints to prevent beads from migrating toward each other. The bionic reconstruction also included agonist-antagonist myoneural interface (AMI) surgery, which surgically reconnects residual agonist and antagonist muscles to restore biological mechanical coupling (Methods). This approach enables proprioceptive feedback and demonstrates enhanced prosthetic control compared to conventional amputations^35,36^. The AMI construct’s large, controlled muscle excursions enable clear differentiation of sensing modality performance. Two AMI muscle pairs were created for each participant: lateral gastrocnemius (GAS) and tibialis anterior (TA) for ankle control, and tibialis posterior (TP) and peroneus longus (PL) for subtalar control (Fig. 3b,d).

**Fig. 3.**
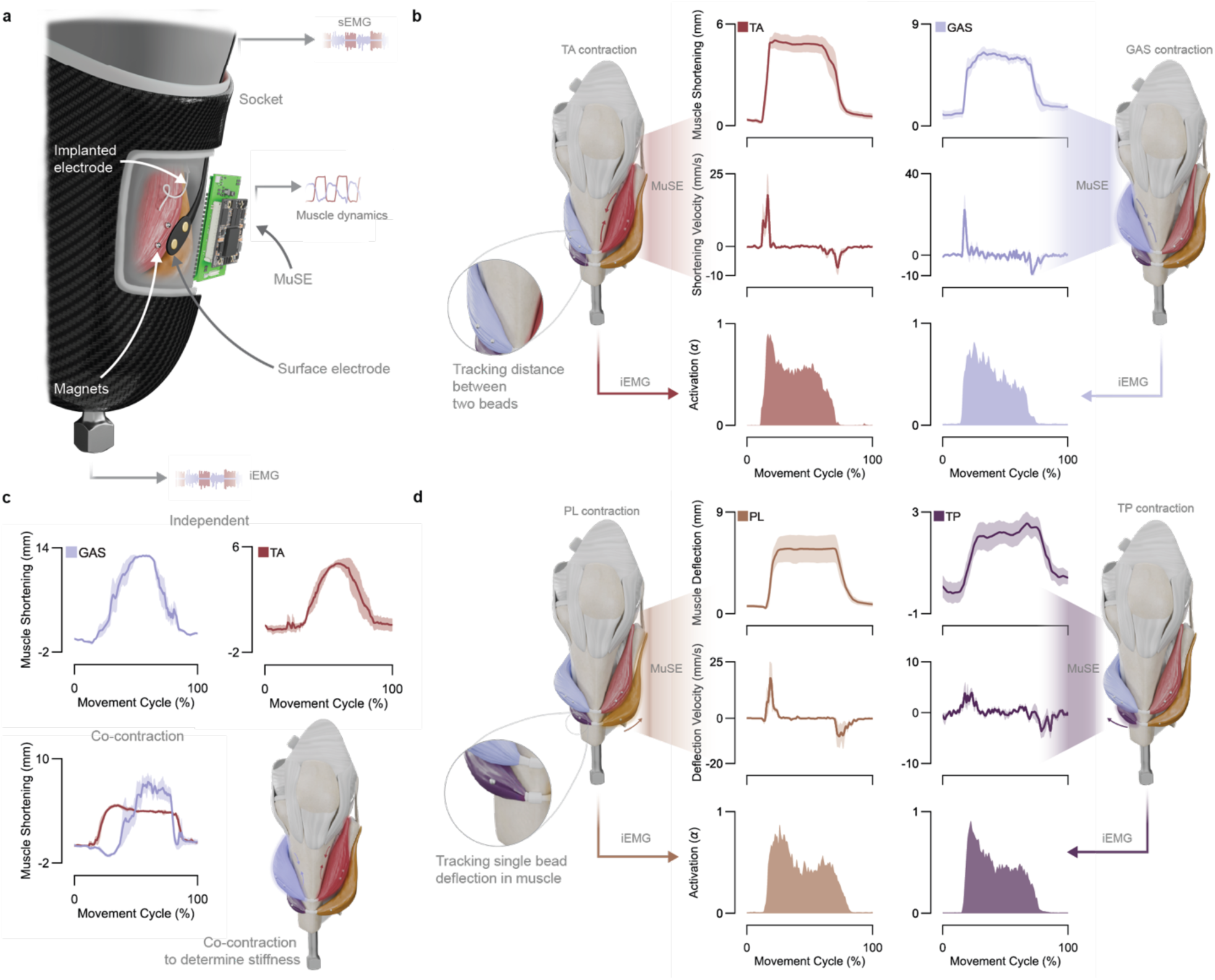
Wireless in vivo continuous sensing of muscle dynamics. **a,** Experimental implementation of all three interfaces – MuSE, iEMG, and sEMG – with residual muscles. Specifically, we implanted magnetic beads and intramuscular electrodes into each muscle and positioned surface electrodes on the skin adjacent each muscle. Subjects wore a prosthetic socket to allow mounting of the sensor boards. iEMG was accessed from a transcutaneous implanted device. MuSE systems tracked bead positions through the socket, liner, and skin. **b,** Evaluation of signal characteristics within the ankle muscles, specifically the TA and GAS muscles with two magnetic beads each. Plots represent average muscle shortening and velocity from the MuSE and muscle activation from iEMG at 100% perceived effort for a 5-second contract-hold-release movement cycle (n = 3 trials per subject, n = 2 subjects). **c,** Muscle deflections from Subject 3 are shown, where the subject was asked to slowly ramp up and then down muscle contraction, first each muscle independently and then both simultaneously, over a 5 second movement cycle (n = 3 trials per subject per movement type, n = 1 subject). **d,** Assessment of signal characteristics within the subtalar muscles, specifically the PL and TP muscles with one magnetic bead each. Plots represent average muscle deflection and velocity from the MuSE and muscle activation from iEMG at 100% perceived effort for a 5-second contract-hold-release movement cycle (n = 3 trials per subject, n = 2 subjects). All muscle deflection and velocity shaded regions indicate SEM.

We simultaneously acquired signals from three sensing platforms: MuSE, implanted electromyography (iEMG), and surface electromyography (sEMG). Participants wore a custom prosthetic socket with up to four MuSE systems positioned over each muscle based on maximum magnetic field strength at palpated muscle locations, with surface electrodes placed at the same anatomical landmarks. All signals were sampled at 1 kHz, with EMG signals processed through band-pass filtering (20-450 Hz), rectification, and envelope extraction (10 Hz cutoff) with bilinear activation dynamics to generate muscle activation signals (Methods). The MuSE raw muscle length signals were low-pass filtered at 10 Hz to capture the physiological bandwidth of muscle movement. The MuSE achieved simultaneous tracking of multiple implanted magnets through the prosthetic socket across tissue depths of 3-6 cm (Fig. 3a, Supplementary Fig. 14). During maximal voluntary contractions, the MuSE revealed mean muscle fascicle shortening of 6.2 mm for GAS and 4.8 mm for TA and mean muscle movement of 5.7 mm for PL and 2.2 mm for TP, with contraction profiles showing rapid onset and sustained isometric phases (Fig. 3b,d). MuSE enabled direct calculation of muscle fascicle shortening velocities from length measurements. The system also allowed subjects to demonstrate muscle contraction ramping and coordinated co-contraction patterns within muscle pairs, indicating potential for impedance modulation control for neuroprosthetic performance (Fig. 3c). We observed fundamental differences in signal characteristics between MuSE and EMG during sustained isometric contractions: while EMG signals exhibited temporal decay throughout the contraction period, the MuSE signals remained stable (Fig. 3b), providing consistent muscle state information throughout sustained contractions. This decay in EMG amplitude does not reflect changes in the underlying isometric muscle state, suggesting the MuSE may have advantages for real-time neuroprosthetic interface applications requiring consistent muscle state information. In addition, no complications occurred from magnetic bead implantation across the three subjects over an average implantation period of more than one year, with no bead migration or associated implant irritation observed (Supplementary Table 2).

### MuSE as a human-machine interface

To evaluate the MuSE system’s ability to work as a neuroprosthetic interface compared to standard EMG methodology, we designed two experiments: (i) a free-space mirroring task to assess the ability to reconstruct single-DOF joint kinematics (Fig. 4a), and (ii) a virtual 2D cursor task to evaluate multi-DOF prosthetic control (Fig. 5a). For the mirroring experiment, subjects were instrumented with a goniometer (Biometrics Ltd., Newport, UK) to capture reference kinematics. A metronome provided audio cues ranging from 60 to 160 BPM in 20 BPM increments, prompting subjects to move between the extremes of their range of motion over 30-second trials (Supplementary Video 3). MuSE, sEMG, and iEMG signals from the residual limb were digitally synchronized with goniometer data sampled at 1 kHz. For EMG-based interfaces, we applied a learnable forward-dynamics model, which provided a second-order description of the ankle joint where muscle activation dynamically modulates joint impedance and position, with parameters optimized to minimize error between predicted and reference kinematics (Supplementary Note 3). For the MuSE, opposing length changes from the TA and GAS muscles were fused into a bidirectional signal representing intended joint angle, with a second-order polynomial optimized to transform muscle length changes to joint space.

**Fig. 4.**
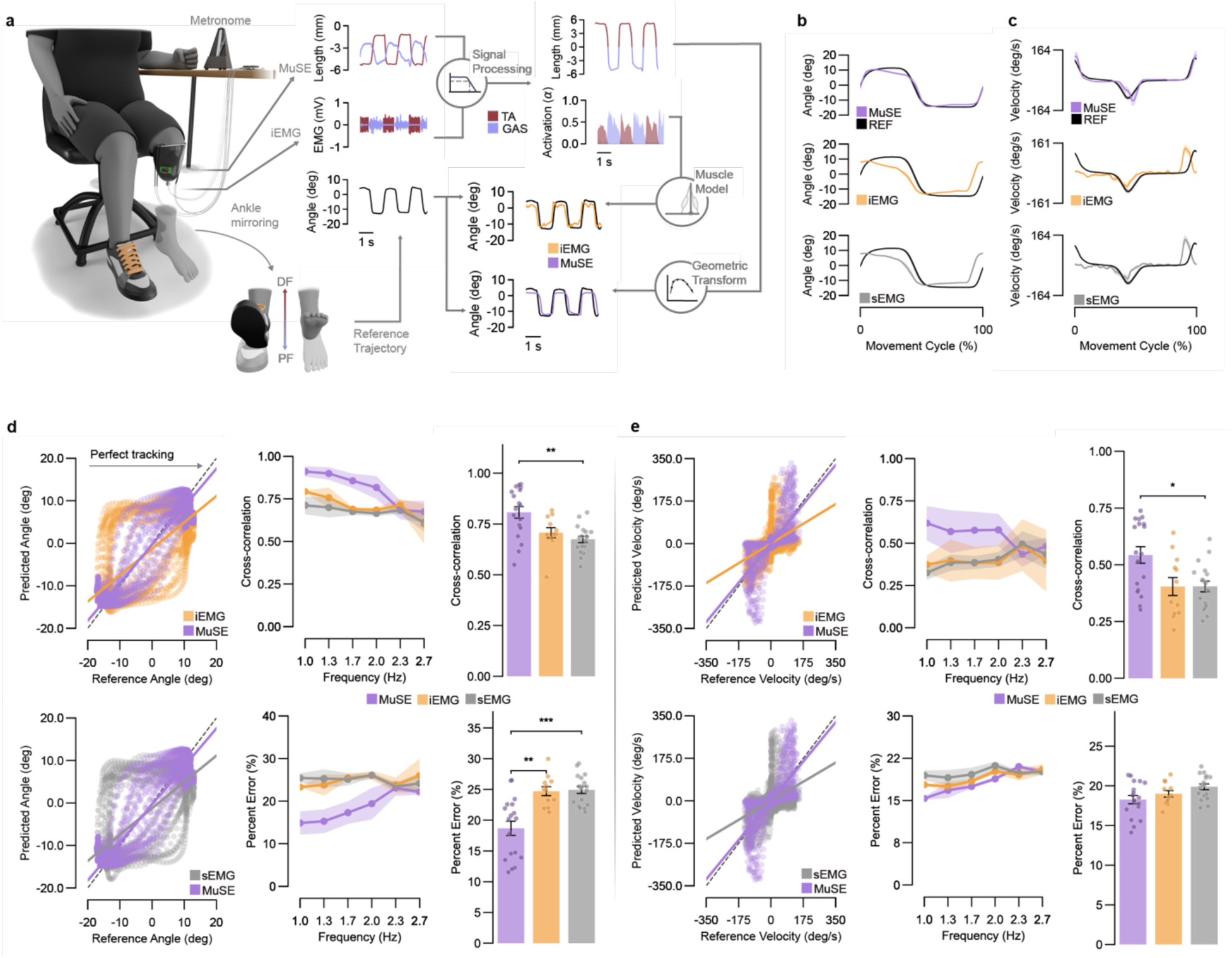
MuSE enables accurate reconstruction of joint kinematics. **a,** Evaluation of mirrored joint kinematics between intact ankle and phantom ankle as the subject tracks the full range of movement at a specified frequency designated through a metronome. Representative plots depict the reference trajectory generated from intact ankle kinematics, while EMG and MuSE signals are collected from the amputated side TA and GAS residual muscles. These signals are processed into activation and combined length signals. The combined length undergoes geometric transformation, while activation signals pass through a bilinear muscle model to generate modeled ankle kinematics for comparison with the reference trajectory. **b,** Representative plot of Subject 2 ankle angle trajectory comparison across all interfaces between reference and modeled kinematics at 1 Hz movement frequency. **c,** Representative plot of Subject 2 ankle velocity trajectory comparison across all interfaces between reference and modeled kinematics at 1 Hz movement frequency. **d,e,** Example scatter profiles of reference angle and velocity plotted against predicted angle and velocity for Subject 2’s trial at 1 Hz, with the dashed line representing perfect tracking between the two. Cross-correlation and percent error for ankle angle **(d)** and velocity **(e)** tracking across each frequency (n = 6 frequencies per subject, n = 3 subjects, n = 18 trials for sEMG and the MuSE, n = 12 for iEMG). Lines of best fit for the MuSE, iEMG, and sEMG are shown. The statistical test to determine significance **(d,e)** was Kruskal–Wallis with post hoc comparisons and Bonferroni correction. ***P < 0.001, **P < 0.01, *P < 0.05. All bars indicate mean and SEM. All shaded regions indicate SEM.

**Fig. 5.**
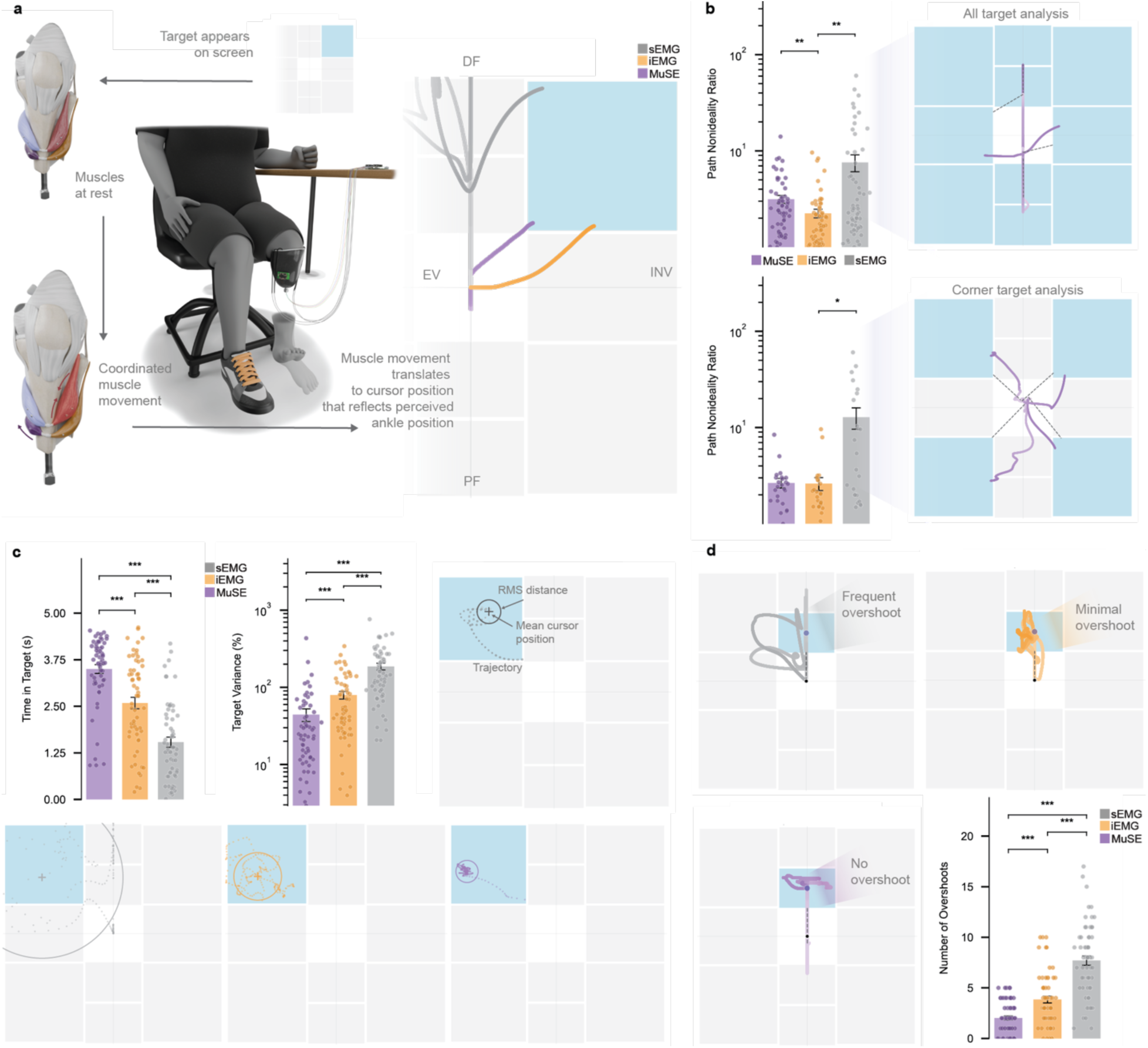
MuSE provides superior neuroprosthetic performance. **a,** Virtual 2D map representing the ankle range of motion, with the horizontal axis representing subtalar motion (left side: eversion (EV); right side: inversion (INV)) and the vertical axis representing ankle motion (up: dorsiflexion (DF); down: plantarflexion (PF)). Representative cursor trajectories from the MuSE, iEMG, and sEMG interfaces to a corner target from Subject 1, requiring simultaneous muscle activation. **b,** Path non-ideality of cursor trajectory to target for all targets together (n = 2 subjects, n = 3 trials per subject, n = 10 independent targets per trial) and corner targets only (n = 2 subjects, n = 3 trials per subject, n = 4 independent targets per trial). **c,** Summary of target reach performance metrics: cursor variance and time in target (n = 2 subjects, n = 3 trials per subject, n = 10 independent targets per trial). Representative plots from Subject 1 demonstrate cursor variance across all interfaces. Dotted lines represent trajectory after target reach (+ indicates cursor position mean and circle indicates root-mean-square of cursor position after target reach). **d,** Summary of target overshoot performance (n = 2 subjects, n = 3 trials per subject, n = 10 independent targets per trial). Statistical tests used Kruskal–Wallis with post hoc comparisons and Bonferroni correction. ***P < 0.001, **P < 0.01, *P < 0.05. All bars indicate mean and SEM.

Representative tracking performance at 1 Hz demonstrated that MuSE signals closely matched reference trajectories, whereas EMG-based reconstruction exhibited larger deviations and noticeable temporal lag (Fig. 4b,c). The MuSE achieved significantly superior angle tracking compared to sEMG, with higher cross-correlation (0.807 ± 0.029 vs 0.674 ± 0.016, **P < 0.01) and lower percent error (18.69 ± 1.17% vs 24.92 ± 0.58%, ***P < 0.001) (Fig. 4d). Velocity tracking demonstrated MuSE’s enhanced temporal resolution, achieving significantly higher cross-correlation compared to sEMG (0.543 ± 0.036 vs 0.405 ± 0.024, *P < 0.05) (Fig. 4e). No significant performance differences were observed between sEMG and iEMG, likely due to both maintaining high muscle selectivity. Scatter plots of predicted versus reference trajectories illustrated these differences, with MuSE data clustering tightly around perfect correlation while EMG-based predictions showed greater dispersion and systematic deviations (Fig. 4d,e). Raw signals from the MuSE demonstrated a waveform that closely approximated the target joint kinematics. EMG signals, even when integrated with an optimized neuromuscular model, failed to track intended joint kinematics as accurately as the MuSE across the tested frequency range. Muscle dynamics signals required only simple geometric scaling from muscle displacement to ankle rotation, directly relating muscle mechanical state to joint position, while the EMG approach with an optimized neuromuscular model could not replicate motion as closely. While EMG necessitates complex filtering to extract meaningful control signals, the MuSE provides direct kinematic information, reducing cumulative processing errors, delay, and loss of information. The system enables increased bandwidth in tracking rapid kinematic changes, maintaining joint information that EMG reconstruction either attenuated or delayed.

To assess multi-DOF prosthetic control, we evaluated interface performance through a virtual 2D cursor task that mapped ankle and subtalar muscle commands to movement in a coordinate system representing physiological joint ranges (Fig. 5a, Supplementary Video 4 and Supplementary Note 4). Subjects performed three sequential tasks: move the cursor into a target window, maintain position for 5 seconds, then return to neutral. For cursor path trajectories, iEMG demonstrated the lowest path non-ideality for all targets combined (2.25 ± 0.23) compared to sEMG (7.59 ± 1.52) and MuSE (3.15 ± 0.30) (**P < 0.01) (Fig. 5b), with similar advantages for corner targets where iEMG (2.62 ± 0.41) significantly outperformed sEMG (12.82 ± 3.28, *P < 0.05). However, the MuSE provided superior control stability, achieving significantly lower cursor variance (44.32 ± 8.29%) and longer time-in-target (3.50 ± 0.12 s) compared to iEMG (79.49 ± 9.15%, 2.59 ± 0.16 s) and sEMG (186.59 ± 18.61%, 1.54 ± 0.14 s) (***P < 0.001) (Fig. 5c). MuSE also demonstrated improved precision with significantly fewer overshoot events (2.02 ± 0.21) compared to iEMG (3.85 ± 0.34, ***P < 0.001) and sEMG (7.70 ± 0.47, ***P < 0.001) (Fig. 5d). This stability advantage reflects fundamental differences in signal characteristics: The MuSE system provides continuous measures of muscle contraction, enabling sustained positioning during isometric contractions with minimal fluctuation, whereas EMG signals exhibit temporal decay and threshold-like behavior that limit fine positional adjustments. The improved selectivity of implanted interfaces compared with sEMG underscores the importance of targeted sensing (Supplementary Fig. 15). While iEMG achieved lower path non-ideality, this benefit required highly invasive electrode placement, whereas the MuSE achieved superior stability metrics through minimally invasive 3 mm implants, demonstrating effective multi-DOF control without the invasiveness of wired electrodes.

### Contactless muscle activation sensing

We used a passive coil in conjunction with the implanted magnets to measure muscle activation through bead vibration. This passive coil can be added to the MuSE system as an additional module. The level of neural activation of a muscle is determined by the firing rate of motor units controlling its fibers^37^. We reasoned that vibrations generated by muscle fibers, which are representative of neural signals, can be detected from changes in the magnetic flux resulting from the corresponding vibrations of implanted magnets. This new technique, which we call magnetovibrometry (MVM), represents a form of contactless mechanomyography^38^ that avoids the impedance artifacts inherent in mechanical contact methods. Previous studies have demonstrated that muscle vibration (mechanomyography) can be measured mechanically^39^ or acoustically^40^. In contrast, MVM provides a contactless alternative with highly linear and repeatable responses across subjects.

The inductive sensor comprised an air-core, multi-layer coil wound with 36-AWG copper wire on a 3D-printed former (inner diameter 8 mm, outer diameter 48 mm, length 16 mm; N = 11,500 turns). The sensor was designed for low-frequency sensitivity to muscle twitch vibrations (<200 Hz), with output fed to a low-noise differential amplifier. For in vivo characterization, the inductive sensor was positioned over muscles targeting a single magnet (Fig. 6a). Raw voltage recordings showed decreasing amplitude and increasing frequency with greater muscle effort (Fig. 6b). Zero-crossing frequencies, computed over fixed time windows, increased with muscle effort, consistent with higher muscle twitch frequencies resulting from increased neural firing rates (Fig. 6c). To compare MVM with EMG, simultaneous measurements were collected from implanted electrodes. Normalized MVM signals closely matched EMG in both amplitude and temporal profile (Fig. 6d). Both participants showed increasing MVM signals with increasing effort. (Fig. 6e). MVM amplitude increased with muscle effort in a pattern similar to EMG (Fig. 6f), with a mean correlation coefficient of 0.7 between the two modalities (Fig. 6g). In agreement with previous findings, we found EMG to be highly linear for most of the range of percent efforts, but highly nonlinear at low levels of muscle activity^41^. Previous mechanomyographic work describes either amplitude or frequency increases with muscle effort, depending on sensing modality^42^. With implanted magnets positioned near muscle fibers, MVM exhibited increasing signal frequency and decreasing amplitude, consistent with force dynamics at higher motor unit firing rates. The observed frequencies aligned with previously reported mechanomyography values^43^.

**Fig. 6.**
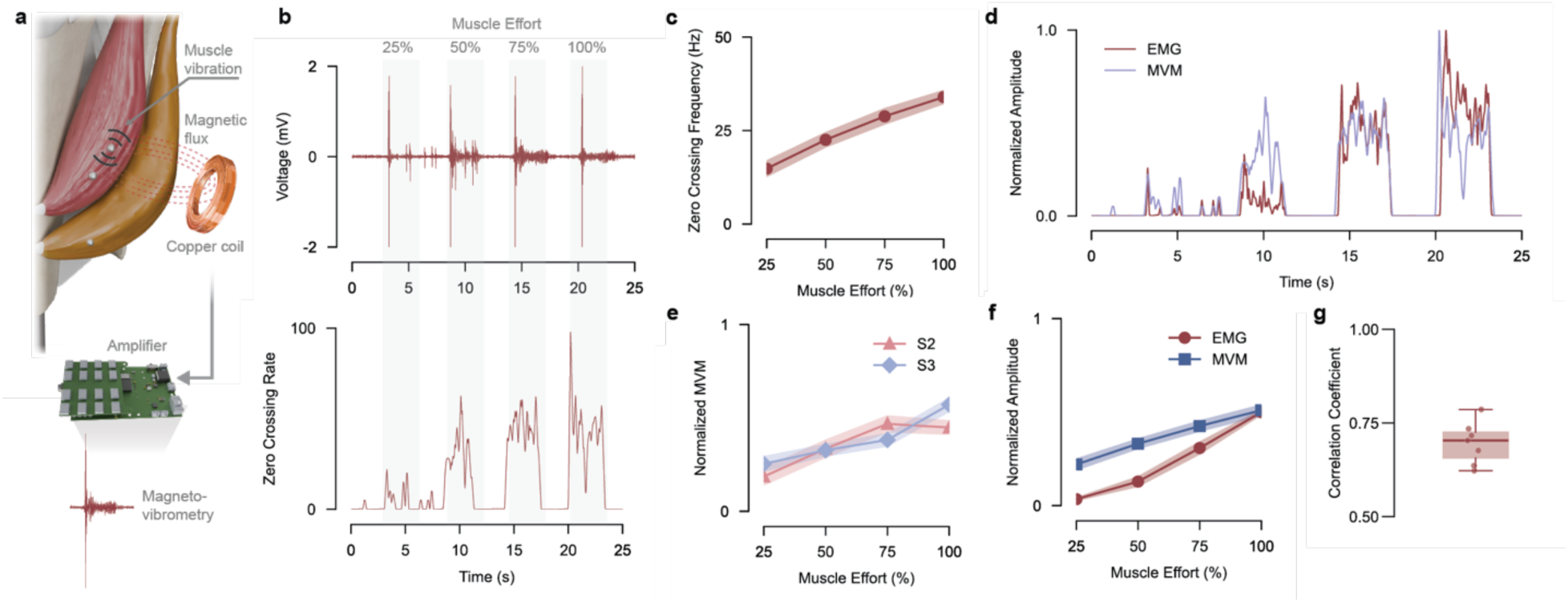
Magnetic detection of muscle activation. **a,** Schematic illustration of the magnetovibrometry (MVM) system, showing inductive coil placement near an implanted permanent magnet to detect magnetic flux changes during muscle contractions. **b,** Representative magnetovibrometry signals show raw voltage measurement corresponding to different muscle effort levels (25%, 50%, 75%, 100% maximum voluntary contraction, indicated by shaded regions). The bottom panel displays the corresponding zero-crossing rate over time after processing the voltage measurement. **c,** Relationship between muscle effort level and zero-crossing frequency, demonstrating a positive correlation between muscle activation and vibration frequency across the physiological effort range. **d,** Time-series comparison of simultaneously recorded EMG and MVM, demonstrating temporal and magnitude correlation between muscle activity measurement modalities. **e,** Normalized MVM signals comparing responses between Subject 2 and Subject 3 across varying muscle effort levels, showing consistent inter-subject patterns. **f,** Quantitative comparison of normalized EMG and MVM amplitude responses across muscle effort levels (25-100%), showing similar increases in both modalities with muscle effort. **g,** Correlation statistics between EMG and MVM measurements across all effort levels demonstrating strong correlation between modalities. Shades represent SEM, 14 trials 7 for each subject.

## Conclusion

The MuSE system demonstrates that real-time, wireless human muscle dynamics can be measured by combining permanently implanted 3 mm diameter magnetic beads with custom electronics, a high-density sensor array and an information-theoretic tracking algorithm. This approach enables simultaneous localization of multiple implanted magnets with sub-millimeter resolution at depths up to 6 cm. We illustrate these capabilities by using the platform to continuously track multiple muscles in amputated human participants and provide intuitive, precise prosthetic control through direct measurement of muscle length and velocity. Unlike EMG and other mechanical sensing approaches that suffer from portability constraints, invasiveness requirements, or imprecision, the MuSE system directly measures the mechanical state of muscle tissue dynamics with minimal surgical invasiveness while providing reliable sub-millimeter measurements. In neuroprosthetic applications, previous systems often scale poorly with additional degrees of freedom due to muscle cross-talk and calibration requirements. In contrast, the MuSE provides joint-specific information directly from muscle mechanics, potentially offering a more scalable and intuitive pathway toward complex human-machine control^44^. In extended testing, we successfully tracked magnetic flux changes with a coil to detect muscle vibrations and applied the signal as a direct mechanical analog to muscle activation.

These advances address several factors that limit tracking fidelity in clinical implementations of magnetic bead localization, including: (i) low sensor density and computationally expensive algorithms that limit spatial resolution and tracking precision, (ii) restricted implantation and sensing depth, since magnetic field strength decreases rapidly as the inverse cube of distance from the sensor, and (iii) superimposed magnetic fields from multiple magnets that complicate state estimation. Beyond the muscle-based sensing for neuroprosthetic control demonstrated in this study, the system has broader potential in human–machine interface domains such as exoskeleton control and rehabilitation monitoring, as well as biomechanical studies in both humans and animals^45^. A key advantage is that passive implants serve as stable, long-term platforms: once implanted, they remain unchanged while new sensors, algorithms, and system integration progressively unlock novel sensing capabilities over time. For instance, by combining magnetomicrometry with magnetovibrometry using the same implants, the MuSE can simultaneously capture muscle length, velocity, and activation information, providing precise, real-time muscle state and force estimation that expand beyond what previous approaches could achieve. The MuSE system’s localization and design principles can extend beyond muscle sensing to applications such as tracking electronics within the gastrointestinal tract^46^ or localizing surgical devices^47^.

Several implementation considerations remain for future development of the MuSE system. First, muscles instrumented with single magnets introduce measurement uncertainty because the system cannot distinguish actual magnet motion from muscle activity versus apparent motion caused by sensor board displacement. This can be mitigated by tracking movement of sensor boards with inertial measurement units (IMUs). Second, high-precision sensing inherently measures total muscle tissue deformation, including both active contractile and passive muscle activity. Biarticular muscles exhibit this complexity, as signals may include passive length changes from proximal joint movement. Although participants maintained constant knee angles to isolate ankle dynamics in this study, future implementations must decouple confounding effects of passive and active muscle movements, such as through fusion of joint and muscle state information^48^. Finally, the large contractile amplitudes observed are partly attributed to the AMI surgical construct, which provides enhanced mechanical strain that amplifies detectable fascicle length changes. Future work should investigate integrating the MuSE system with other surgical neuroprosthetic interfaces to assess performance across different anatomical configurations.

## Methods

### MuSE sensor electronics

The muscle state estimator electronics architecture consisted of three components: (i) a custom sensor board, (ii) a custom carrier board, and (iii) an FPGA system-on-module (SoM). The sensor board contained 640 magnetometers (LIS3MDL, STMicroelectronics) that used serial peripheral interface (SPI) communication protocol. The array is organized as 32 SPI columns, each column serving 20 sensors that share the data lines. Four SPI clocks are evenly distributed across the 32 columns (8 columns per clock). The rows are organized by a cascaded shift-register chain (74LVC594ABQ,115, Nexperia) that drives 20 chip-select (CS) lines. A sensor is uniquely addressed at the intersection of one SPI column (data bus) and one active CS row.

The primary challenge in this high-density sensor configuration is clock distribution: each of the four clock lines must drive 160 LIS3MDL magnetometers (8 columns × 20 rows), presenting a high cumulative capacitive load. We implemented high-current complementary push-pull drivers using discrete MOSFETs (SSM6L56FE,LM, Toshiba) capable of sourcing and sinking high instantaneous current to actively drive both rising and falling clock edges. The signal path for each clock line begins with a voltage level step-up from 1.8 V FPGA logic to 3.3 V sensor logic levels via dual-channel voltage translators (SN74AVC2T244DQMR, Texas Instruments), which directly drive the gates of complementary MOSFET pairs. The complementary push-pull output stage delivers rail-to-rail swing with low resistance to maintain signal integrity even under maximum capacitive load. With clock distribution traces spanning across the board, transmission line effects become significant. Each clock line incorporates series termination resistors positioned near the driver output to match the controlled-impedance PCB traces, damping reflections and minimizing overshoot. Shunt capacitors filter high frequency switching noise and reduce electromagnetic interference. Clock traces are length-matched to maintain synchronization across all sensors sharing a clock line, with continuous ground plane beneath the traces ensuring low-impedance return paths.

Sensor array scanning employs row-sequential addressing: an address is serially loaded into cascaded shift registers to activate one chip select line, enabling all 32 sensors in that row. The FPGA then sequentially polls each of the 32 SPI buses, transmitting read commands on the output data line and receiving three-axis magnetic field data on the input data line. Only the sensor at the intersection of the active chip selects and addressed SPI bus responds.

### MuSE computing electronics and packaging

The carrier board hosts the FPGA SoM (Mercury XU5, Enclustra), power management, timing, and I/O needed to operate the 640 sensor array. A 12 V input drives board power, a voltage buck converter (TPS564208DDCT, Texas Instruments) outputs 3.3 V for sensor power supply. High-speed digital interfaces are routed from the FPGA to the sensor board as the carrier board exposes the 32 parallel SPI channels and 1.8 V logic lines. Ethernet is provided for wired communication, and a WiFi module (LBEE5KL1DX, Murata) provides wireless communication at 2.4 GHz through a flexible antenna (FXP830, Taoglas). A 100 MHz oscillator (ASEMB-100.000MHZ-LY-T, Abracon) supplies a stable timing reference for the SoM. A custom-designed aluminum heatsink and 12 V fan (RS5010B12L-FA, Runda) were added to thermally regulate the FPGA chip. A thin 1 mm layer of copper pads was placed on top of the sensors to allow the sensors to conduct heat away and prevent thermal drift. The sensor board and carrier were connected through either a flex-PCB connector or rigid connector, while the FPGA mounted to the carrier through rigid connectors. An enclosure was 3D-printed to protect the electronics, with final dimensions of 70 mm x 100 mm x 48 mm.

### Information-based tracking algorithm

Based on the dipole tracking principles of Taylor *et al.*^34^, we aimed to solve the inverse problem of estimating magnet position and orientation from the array of 3D magnetic field measurements. In addition to the magnet state, the uniform disturbance field state was estimated, which in most cases is the geomagnetic field. This was done with an extended information filter (INFO) framework. The information filter represents system state through an information matrix 𝐘_0_(inverse covariance 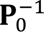) and information vector 𝐲_0_ (inverse covariance times state vector 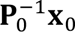), enabling efficient processing of systems with significantly more measurements than states. The INFO operates with an information matrix 𝐘 and information vector 𝐲, initialized as:

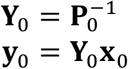

where 𝐏_0_ is the initial covariance matrix and 𝐱_0_ is the initial state estimate.

The algorithm computes information increments from each sensor measurement as 𝐇^𝑇^𝐑^−1^ (measurement innovation), where 𝐇 is the measurement Jacobian and 𝐑 is the measurement noise covariance. For each measurement update, the information matrix and vector are updated as:

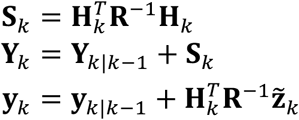

where 𝐒_𝑘_ represents the innovation information contributed by the new measurement. The optimal state estimate is recovered through:

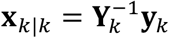

### Magnet tracking simulation and benchtop validation

To evaluate different tracking algorithms, sensor configurations, and accuracy metrics, a virtual environment was constructed to validate the influence of these parameters on state estimation. The simulation framework reproduced sensor measurements by generating expected fields from inputting true parameters of the magnet state and adding Gaussian white noise representative of the sensor noise.

All simulation trials used modeled sensors with 2.5 μT mean noise and tracked magnetic beads with dipole strength of 1.39 nT·m³, matching implanted bead strength. Static and dynamic trials included a uniform geomagnetic disturbance field with magnitude resembling physical values (−20 μT x-direction, 20 μT y-direction, -80 μT z-direction). Sensors were simulated at 1 kHz sampling rate with geometry matching the physical sensor board. Comparative analysis between Levenberg-Marquardt (LM) and extended information filter (INFO) algorithms used identical analytical dipole field models and Jacobian equations.

We conducted multiple simulation experiments using our base hardware configuration of 640 magnetometers. For single magnet static trials, we evaluated tracking performance across 44 spatial configurations, testing four depths (30, 40, 50, 60 mm) from the sensor array with 11 x-y positions per depth generated from a 5×5 arrangement of x-coordinates (0, 10, 20, 30, 40 mm) and y-coordinates (0, 8, 16, 24, 32 mm). For sensor density analysis, we tested five configurations with systematically reduced active sensor counts while maintaining spatial distribution: 60, 120, 196, 320, and 640 active sensors, with a single simulated magnet positioned at (0, 0, 60) mm.

Static simulation experiments used identical algorithmic parameters and protocols. For single magnet trials, each configuration underwent 2 independent trials, while density analysis and magnet pair used 10 independent trials per configuration. All trials employed identical random initial parameter estimates for both algorithms to enable direct comparison, with initial conditions randomly generated within the sensor coverage volume. If either algorithm diverged due to the non-convexity of dipole modeling, another initial guess was initialized for that trial. Each algorithm performed 4,000 estimation iterations (4 simulated seconds), with final performance metrics calculated from the last 500 iterations to ensure steady-state convergence.

For simulated dynamic trials, we assessed real-time tracking capability during continuous magnet motion using pre-computed trajectories, where we allowed both algorithms to first reach steady-state convergence of the initial state before tracking the dynamic movement of either one or two magnets.

Performance across each trial was quantified using mean absolute error (MAE):

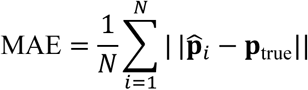

and positional error variance:

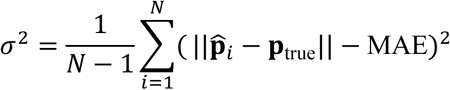

where 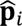 represents estimated position or distance, 𝐩_true_ is true position or distance, and 𝑁 = 500 steady-state iterations.

For benchtop testing with the physical magnet tracking embedded system, two magnets were positioned at [±20, 0, z] mm, spaced 40 mm apart, across four depths (30–60 mm). Both tracking algorithms were implemented in a real-time C++ framework and compared algorithm latencies and estimation errors by analyzing the final 500 iterations at steady-state convergence.

### Clinical study design

This study was conducted under registered clinical trial NCT06391697, which aimed to investigate the clinical safety and efficacy of the magnetic bead tracking system in conjunction with the implanted electrode system (e-OPRA, Integrum AB). The study protocol and implants underwent FDA IDE approval as well as institutional review board approval at both MIT and BWH, where experiments were completed under supervision. Both systems were implanted at the transtibial amputation level in combination with the agonist-antagonist myoneural interface (AMI) bionic reconstruction (Supplementary Note 5).

We recruited 3 participants, two of which had undergone an acute transtibial amputation that received both magnetic bead and implanted electrode implantation, and one of which had a pre-existing amputation with revision to add the magnetic bead tracking system and reimplantation of the implanted electrode system. This subject was previously part of a related study NCT05249049, which investigated the implanted electrode system with a robotic prosthesis. All surgeries and pre-screening to determine eligibility were conducted at Mass General Brigham Hospital (Boston, Massachusetts) under Mass General Brigham IRB protocol #2024P000006. Consent for both surgery and human subjects testing fell under the Mass General Brigham protocol, of which the MIT IRB ceded to. Before surgery and testing, subjects provided their written consent at both MIT and MGB sites. Exclusion criteria for the study included active skin disease, advanced muscle atrophy, or compromised soft tissue in the operative limb; severe comorbidities such as diabetes, peripheral vascular disease, neuropathy, severe phantom pain, or osteoporosis; known future MRI requirements; device allergies; and active smoking or pregnancy. A full detailed list of both inclusion and exclusion criteria and interventions can be found at NCT06391697. The cohort of 3 participants underwent repeated measures of tasks, and the period of recruitment and data collection within this study occurred from 2023 to 2025.

### EMG and MuSE implementation

To simultaneously capture data from all three neural interfaces (MuSE, surface electromyography (sEMG), and implanted electromyography (iEMG)), all measurement hardware was connected concurrently. We utilized the EMG acquisition system developed by Yeon *et al.*^49^, connecting both implanted electrodes from the osseointegrated connector and surface electrodes to the system for each muscle, utilizing four bipolar channels per interface.

For MuSE system positioning, participants wore a custom socket with an opening to accommodate the osseointegrated implant, enabling static mounting of tracking hardware over target muscles. Surface electrodes utilized the same flexible, thin-film design developed by Yeon *et al.*^49^ to capture sEMG signals within the socket. Surface electrode placement was guided by palpation of known AMI construct locations, with electrodes positioned directly over these anatomical landmarks. To determine optimal MuSE board placement across the socket surface, we scanned each palpated AMI construct region using the array of magnetic field sensors. For each muscle, we identified locations containing maximum magnetic field strength, whether from magnet pairs or single magnets, and marked these positions before adhering the boards to the socket. In most configurations, we allocated one dedicated board per muscle, positioning up to four boards around the socket perimeter for up to four muscles. Board outputs consisted of either distance measurements between magnet pairs or, for muscles with single magnets, deflection measurements as the muscle underwent volitional activation.

### Free-space mirroring task

To evaluate how each interface can predict intended joint kinematics, we designed a free-space mirroring task similar to Shu *et al*^50^. To generate a reference trajectory, subjects were instructed to mirror intact ankle kinematics with their perceived phantom ankle motion while we measured MuSE, sEMG, and iEMG signals on the residual limb. Subjects sat on a stool to ensure full range of motion could be achieved, and a metronome track with tempo ranging from 60 to 160 BPM in 20 BPM increments was provided as an audio cue. The metronome indicated on each beat to move to one end of the range of motion (for example, from dorsiflexion to plantarflexion) over the course of 30 seconds as the beat played. The intact ankle was instrumented with a goniometer (Biometrics Ltd., Newport, UK) to capture ankle kinematics. The neural signals and goniometer were digitally synced to ensure correct time alignment for data post-processing and zeroed at ankle rest position of 90 degrees. Angular data was collected at 1 kHz, and a fourth-order Bessel low-pass filter (cutoff frequency: 10 Hz) was applied. For each trial, subjects had to perform the mirroring for 30 seconds. To model the intended ankle kinematics of the phantom limb from muscle activation collected by iEMG and sEMG, we applied a learnable, forward-dynamics model of a single-DOF joint actuated by a muscle pair. We developed a second-order description of the ankle joint where muscle activation input dynamically modulated the joint’s impedance and position. The model’s parameters were optimized to minimize the mean squared error between the experimentally observed reference and modeled kinematics.

To create a direct kinematic estimate from the MuSE, the two length signals from the TA and GAS muscles were combined by fusing the opposing length changes into a single, bidirectional signal representing the intended joint angle. To determine the geometric transformation between the muscle length changes to joint space, a second-order polynomial was optimized to minimize the mean squared error between the MuSE signal and the intact joint angle.

The primary analysis compared the predictive accuracy of the direct MuSE-based estimate against the EMG-driven forward-dynamics model. Both predictions were benchmarked against the ground-truth goniometer data. This comparison was performed for both sEMG and iEMG inputs across all metronome tempos from the 3 subjects. To quantify performance, we evaluated both the predicted joint angle and joint angular velocity. Evaluated metrics included the percent error difference between reference and predicted values. Cross-correlation was also computed to assess the phase relationship between the predicted and reference trajectories. This dual analysis of position and velocity provided a comprehensive assessment of each interface’s ability to decode the user’s intended kinematics.

### 2D virtual GUI task

Outcome measures for evaluating neuroprosthetic control performance were derived from free-space control experiments conducted with visual feedback with similar experimental design as Clites *et al*^36^. Subjects controlled a virtual prosthetic joint position represented as a cursor across a 2D GUI. During volitional control tasks, we assessed each participant’s ability to independently modulate prosthetic joint movement using either a sEMG, iEMG or MuSE controller. Real-time cursor position was visualized on a computer display as locations in two-dimensional joint space, with plantar flexion-dorsiflexion represented on the vertical axis and inversion-eversion on the horizontal axis.

Prior to experimental data collection, activation scaling parameters were subsequently fine-tuned during a calibration phase where participants practiced reaching targets on the GUI with visual feedback and confirmed they could adequately reach each target. During this calibration, participants provided feedback on movement sensitivity, with requests for increased or decreased responsiveness leading to tuning the scaling parameters.

While observing the graphical interface, participants performed three sequential volitional control tasks: (i) move the cursor to within a predetermined rectangular target window in joint angle space; (ii) maintain the cursor within this target window for 5 seconds; and (iii) return the cursor to neutral rest position. Following familiarization with the experimental paradigm, participants repeatedly performed the task at 10 randomly ordered locations across the full virtual prosthetic joint angle space. Neuroprosthetic controllers were tested sequentially in randomized order to minimize fatigue effects. Each participant had 3 trials for each interface.

Performance was quantified using the following metrics: Path nonideality was defined as the angle-space distance traversed during initial movement from rest to target position (task i), normalized to the ideal distance between cursor position at rest and target entry box, with lower scores indicating improved performance. Time in target measured the duration participants maintained the cursor position within the target window during the 5-second hold task (task ii), with higher scores reflecting better performance.

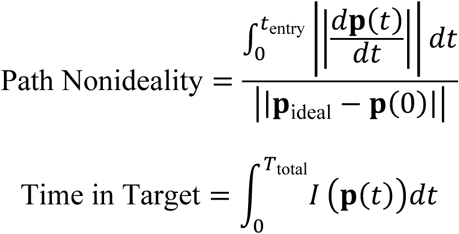

where 𝐩(𝑡) represents cursor trajectory, 𝑡_entry_ is time of first target entry, 𝐩_ideal_ is the closest point on target boundary to starting position 𝐩(0), 𝑇_total_ is total movement duration, 𝐼(𝐩(𝑡)) is an indicator function equal to 1 when cursor is within target and 0 otherwise,

Performance across each trial was additionally quantified using:

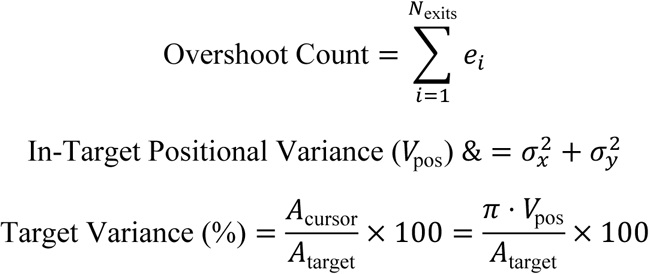

where 𝑒_𝑖_ represents individual target exits, and 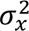 and 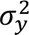 are the positional variances in the x and y directions for in-target positions. 𝐴_cursor_ is the area of cursor variance, while 𝐴_target_ is the area of the target.

### Magnetovibrometry

Sensor inductance was estimated with Wheeler’s multilayer formula L (µH) = (0.8 · r_1_² · N²) / (6r_1_ + 9l + 10*(r_2_-r_1_)), where r_1_ is the radius of the inner diameter, r_2_ is radius of outer diameter, l is the length of the coil and N is number of turns, yielding an inductance of 1.91 H. Sensor output was fed to a low-noise differential instrumentation amplifier (AD8422BRMZ, Analog Devices) then to an analog-to-digital converter (ADS1299, Texas Instruments) similar to the EMG electronics. For human testing, the coil was taped to the skin using medical tape over the proximal TA magnet. Participants produced 25, 50, 75, and 100% MVC, yielding 14 trials across 2 subjects with simultaneous implanted EMG recordings. MVM signals were filtered with a 60 Hz notch and a 10-150 Hz Bessel bandpass filter. Segments were defined by velocity-spike triggers (first-derivative threshold) into MVC effort windows. Zero crossing rate was computed across the whole signal and by segments using a 200 ms sliding window with amplitude gating to exclude baseline noise.

### Statistics

Statistical analyses were performed using Python statistical analysis packages such as numpy and scipy. Data are presented as mean ± standard error of the mean (SEM) unless otherwise specified. Statistical significance was set at α = 0.05, with Bonferroni correction applied for multiple comparisons. Prior to statistical testing, data distributions were assessed using the Shapiro-Wilk test for normality. For multiple group comparisons, Kruskal-Wallis tests were performed to assess overall differences between groups, followed by post hoc pairwise comparisons using Wilcoxon rank-sum test with Bonferroni correction for multiple comparisons.

## Acknowledgment

Funding was provided through the K. Lisa Yang Center for Bionics at MIT in addition to the MIT Media Lab Consortia. C.C.S is supported by the NSF Graduate Research Fellowship. The authors thank Curtis Nelson and Andrew Ferencz for their extensive consultation and dedication on both hardware and FPGA embedded system development. We extend our gratitude to our clinical collaborators Kendall Clites, Rachael Chiao, Grace Cason, and Corey Sullivan at Brigham and Women’s Hospital. We also thank Izaiaha Hayes for his artistic contributions.

## Author contributions

C.C.S., C.R.T., R.J.C., and H.M.H. conceived the MuSE device concept. C.C.S., C.R.T., S.H.Y, R.J.C., D.O., G.H., T.S., J.Q., M.J.C., and H.M.H. contributed to methodology, specifically for the clinical implementation of the MuSE system in humans. M.J.C. conducted the surgeries and appropriate clinical follow up. C.C.S., C.R.T., S.H.Y, R.J.C., and H.M.H. performed electronic system design and implementation of the MuSE and INFO tracking algorithm. C.C.S., S.H.Y and D.O. developed the simulation environment for evaluating magnet tracking. C.C.S., C.R.T., R.J.C., D.O., G.H., J.Q., J.X., D.L., A.R., and S.B. contributed to experimental investigation and data collection of the MuSE for humans. C.C.S., C.R.T., D.O. and G.H. analyzed data and prepared the main figures. C.R.T., S.H.Y., A.L., E.G.C., J.A.P., and H.M.H. devised and validated the magnetometer calibration process. C.C.S. and C.R.T. wrote the original manuscript and C.C.S., C.R.T., R.J.C., T.S., and H.M.H. edited the manuscript. H.M.H. provided supervision and funding for the research. All authors revised and approved the final manuscript.

## Data availability

All data are available in the main text or the supplementary materials.

## Code availability

Simulation framework to evaluate tracking algorithms in Python is available on GitHub. Algorithm implementation of the information filter (INFO) in Python is on GitHub.

