## Additional supplementary material for "A wireless magnetic implant system for continuous neuromuscular sensing"

#### Supplementary Information

### Table of Contents

**Supplementary Note 1: Magnetometer array calibration**

**Supplementary Note 2: Information filter description and implementation**

**Supplementary Note 3: Biophysical model for kinematic reconstruction**

**Supplementary Note 4: Neuromuscular controller**

**Supplementary Note 5: Surgical details and implementation**

**Figure S1. Schematic of sensor logic**

**Figure S2. Schematic of clock driver circuit**

**Figure S3. Magnetometer calibration**

**Figure S4. Benchtop setup to validate magnetometer calibration**

**Figure S5. Calibration process**

**Figure S6. Magnetometer orientation calibration of four magnetometer arrays**

**Figure S7. Repeatability of the magnetometer array orientation calibration**

**Figure S8. Translation calibration and repeatability**

**Figure S9. Static error of tracking before and after adding pose information**

**Figure S10. Inter-magnet distance tracking errors before and after adding pose information**

**Figure S11. Magnetic field sensor noise dynamics in steady-state**

**Figure S12. Single magnet simulation variance**

**Figure S13. Magnetic field strength with increasing distance from magnet**

**Figure S14. Tracking depth of magnet within the human body**

**Figure S15. Muscle cross-talk example in GUI task**

**Table S1. Comparison with other magnetic-based sensing devices.**

**Table S2. Study population characteristics**

**Caption for Supplementary Video 1**

**Caption for Supplementary Video 2**

**Caption for Supplementary Video 3**

**Caption for Supplementary Video 4**

**References**

#### Supplementary Note 1: Magnetometer array calibration

To determine magnetometer array orientations and positions, we used two point-cloud registration techniques one after the other, one to perform the orientation calibration and one to perform the translation calibration. Before calibration, the magnetometer arrays each have their own coordinate frame, and after the magnetometer array poses calibration, they all share a single global frame (Fig. S3). The orientation calibration is performed using the Kabsch algorithm to align the ambient magnetic field point clouds collected during the hard- and soft-iron calibration. The position calibration is performed by aligning the simultaneously-tracked path of a calibration magnet as independently tracked by each magnetometer array. We validate our method using repeatability testing in the context of a benchtop test with four 96-sensor magnetometer arrays.

To validate our magnetometer pose calibration process, we developed a benchtop test setup (Fig. S4) to perform the pose calibration and validated its use specifically in the context of tracking four pairs of magnets, each pair being inserted into one muscle in the residual limb of a patient, with four magnetometer arrays outside the residual limb to track the magnet pair distances. To do this, we created a physical model of a residual limb by performing an MRI scan of a patient's residual limb. We then 3D-printed the bone geometry and formed a silicone model of the soft tissue around the printed bone geometry.

We chose locations for the magnets at locations corresponding to the lateral gastrocnemius, tibialis anterior, tibialis posterior, and peroneus longus at depths of approximately 10-15 mm. We affixed four magnet pairs with a pair-separation-distance of approximately 35 mm to acrylic bars via press-fitting, then inserted these acrylic bars into the residual model at these estimated depths and insertion sites. We then created a thermoplastic socket fit to the residual limb model and attached four magnetometer arrays to the outside of the socket. To account for the contours on the outside of the socket, we used moldable silicone to create flat mounting surfaces for the magnetometer arrays. We affixed the arrays using double-sided adhesive, with each array as close as possible to its corresponding magnet pair. Each magnetometer array was equipped with an 8x12 array of three-axis tunneling-magnetoresistive magnetometer integrated circuits (LIS3MDL, STMicroelectronics) in a grid with a 5.08 mm spacing, all sensing at 155 Hz and communicating with a laptop computer via a custom microcontroller interface printed circuit board assembly.

To compensate for hard- and soft-iron offsets, we use the standard ellipsoid-fitting calibration strategy to accurately calibrate the magnetometers' sensing. Specifically, we first rotate the rigidly-affixed magnetometers together in an approximately spatially-uniform field (keeping them away from anything magnetic or ferromagnetic) to collect an initial calibration data set. We then perform linearized algebraic ellipsoid fits on this data to get the calibrated magnetic fields,

$$\bar{\mathbf{B}}_i = g_i \mathbf{Z}_i (\mathbf{B}_i - \mathbf{v}_i),$$

where, for the $i$th magnetometer, $\mathbf{B}_i$ is the raw magnetic field measurement, $\mathbf{v}_i$ is the hard-iron offsets vector, $\mathbf{Z}_i$ is the soft iron correction matrix, and $g_i$ is the relative scaling factor. This strategy accounts for permanent and temporary magnetization (hard-iron effects) of and magnetic field redirection (soft-iron effects) by the onboard components of the magnetometer array electronics, but it doesn't account for relative orientation differences between magnetometers. Thus, new techniques were needed to account for variation in the magnetometer orientations as well as to determine the positions of magnetometer arrays relative to one another.

The magnetometer array poses calibration is a multi-step process (Fig. S5). First, we calibrate the magnetometers to remove hard and soft-iron effects and to scale their gains relative to one another. Next, we re-use the dataset collected from the traditional calibration to find the relative orientations of all the magnetometers on each array and to determine a bias orientation to correct orientations offsets for the reference sensor of each array. We then repeat this strategy to find the orientations of all reference sensors relative to each other to determine the global orientations of each array. Finally, to determine array positions, we track a reference magnet from each array locally and use the tracked paths of the reference magnet to determine the global positions of each array.

To account for the orientation differences between magnetometers, we perform a pure-rotation point cloud registration on the calibrated magnetic fields, $\bar{\mathbf{B}}$, using the Kabsch algorithm. For each magnetometer (the $i$th magnetometer) of each array (the $k$th array), we compute the cross-covariance matrix between the calibrated measurements, $\bar{\mathbf{B}}_{ki}$, of that magnetometer and the calibrated measurements, $\bar{\mathbf{B}}_{k\hat{\imath}}$, of a most-central magnetometer (the $\hat{\imath}$th magnetometer) on that array, where each set of magnetic field measurements is of dimensions 3x$N$. We then deconstruct this cross-covariance matrix into its singular value decomposition,

$$\bar{\mathbf{B}}_{ki}\bar{\mathbf{B}}_{k\hat{\imath}}^{\mathsf{T}} = \mathbf{U\Sigma V}^{\mathsf{T}}$$

and compute the local rotation matrix as

$$\bar{\mathbf{L}}_{ki} = \mathbf{V}\begin{bmatrix} 1 & 0 & 0 \\ 0 & 1 & 0 \\ 0 & 0 & \det(\mathbf{VU}^{\mathsf{T}}) \end{bmatrix}\mathbf{U}^{\mathsf{T}}.$$

Noting that in our specific application here we expect all magnetometers on a given array to be aligned roughly in the same direction as the array and for manufacturing and assembly misalignments to be symmetrically distributed about that alignment, we then here compute the bias orientation adjustment $\mathbf{A}_k$ of the data as the rotation matrix that minimizes the summed magnitudes of the rotations as

$$\underset{\mathbf{A}_k(\theta_k,\phi_k,\psi_k)}{\arg\min} \sum_{i=1}^{N} \Theta\left(\mathbf{A}_k\bar{\mathbf{L}}_{ki}\right),$$

where $\Theta(\cdot)$ represents the magnitude of the rotation angle of the rotation matrix product and $\mathbf{A}_k(\theta_k, \phi_k, \psi_k)$ represents a rotation matrix with roll $\theta_k$, pitch $\phi_k$, and yaw $\psi_k$. This is the rotation matrix from the reference magnetometer orientation to the "middle" of all magnetometer orientations, for each array. We then update all local rotation matrices for the $k$th array as

$$\mathbf{L}_{ki} = \mathbf{A}_k\bar{\mathbf{L}}_{ki}$$

We next wish to determine whether this orientations calibration is sufficiently accurate to compensate for manufacturing and assembly variability. We split the full dataset into three subsets and repeat the local orientations calibration process on each subset. Comparing with the orientation results from the full dataset, we compute the angular error for each magnetometer for each data subset as $\Theta(\mathbf{L}_{ki}^{\mathsf{T}}\mathbf{L}_{ki}^{(1)})$, $\Theta(\mathbf{L}_{ki}^{\mathsf{T}}\mathbf{L}_{ki}^{(2)})$, and $\Theta(\mathbf{L}_{ki}^{\mathsf{T}}\mathbf{L}_{ki}^{(3)})$, and we compute the root-mean-square of these three angular errors for each magnetometer, determining the distribution of RMS angular errors for each array. We then compare this distribution with the distribution of all orientation magnitudes, $\Theta(\mathbf{L}_i)$, for each array. As a rule of thumb, if the median of the RMS angular errors is less than the median of the orientation magnitudes, we deem the calibration to have been successful.

To determine global array orientations, we now choose a most-central magnetometer, $\hat{\imath}$, from each array, and we repeat the process above to compute the global rotation $\mathbf{R}_k$ from the local-bias-adjusted most-central-magnetometer measurements, $\mathbf{L}_{k\hat{\imath}}\bar{\mathbf{B}}_{k\hat{\imath}}$, of each array to the local-bias-adjusted most-central-magnetometer measurements of the first array, $\mathbf{L}_{1\hat{\imath}}\bar{\mathbf{B}}_{1\hat{\imath}}$. Upon computing these global orientations, $\mathbf{R}_k$, for each array, we then transform future magnetic field measurements into the global frame as

$$\tilde{\mathbf{B}}_{ki} = \mathbf{R}_k \mathbf{L}_{ki} \bar{\mathbf{B}}_{ki}.$$

As a check to the calibration, we also apply this transformation to the calibration data to visually observe its alignment.

At this point, we know the orientations of all magnetometer arrays and their constituent magnetometers in the global frame, but we do not yet know the positions of these arrays in the global frame. To determine these positions, we proceed to a separate calibration strategy.

**Position Calibration**

To account for relative positions of the magnetometer arrays, we next perform a pure-*translation* point cloud registration on the tracked path of a reference magnet seen by all magnetometer arrays. Specifically, we fix the magnetometer arrays (already fixed relative to one another) relative to the ambient magnetic field, and we zero out the ambient field disturbance. We manually introduce a single reference magnet (8 mm N52 sphere, SM Magnetics, SP100352) with a spiraling path that can be seen by all magnetometer arrays, and we track this reference magnet from each array separately in its own local frame using locally-oriented magnetic field measurements, then rotate the tracked magnet locations into the global frame via the global rotation matrix $\mathbf{R}_k$. To ensure that noisy data does not adversely affect the calibration, we then remove all tracked points where the magnet was far away (>100 mm) from the center of any magnetometer array. We then apply a translation-only rigid body transformation to the tracked magnet locations, bringing all magnetometer arrays into a unified reference frame, by calculating the centroids of each point cloud over all time points, $t$, as

$$\mathbf{p}_k = \left[ \frac{1}{N} \sum_t x_{kt} \quad \frac{1}{N} \sum_t y_{kt} \quad \frac{1}{N} \sum_t z_{kt} \right]^\top,$$

and transforming all magnetometer positions into the global frame as

$$\mathbf{s}_{ki}^{\text{global}} = \mathbf{R}_k \mathbf{s}_{ki}^{\text{local}} + (\mathbf{p}_1 - \mathbf{p}_k),$$

where $\mathbf{s}_{ki} = (s_{kix}, s_{kiy}, s_{kiz})^\top$ is the position of the $i$th magnetometer of the $k$th magnetometer array.

To determine the repeatability of this array position calibration, we then split the dataset into three subsets, re-computed the position offsets from each, and calculated the standard deviations of these offsets for each array and axis.

Once we have the global positions of all magnetometers, only the magnetic field measurements need to be transformed to determine the three-axis magnetic fields at each position in the global frame. In the specific application of magnet tracking, the tracking algorithm is then performed with all magnet state variables being in the global frame.

If the initial estimates of the magnet poses are in the local frame, these can be transformed into the global frame as

$$\begin{aligned} \mathbf{x}_j^{\text{global}} &= \mathbf{R}_k \mathbf{x}_j^{\text{local}} + (\mathbf{p}_1 - \mathbf{p}_k) \\ \mathbf{m}_j^{\text{global}} &= \mathbf{R}_k \mathbf{m}_j^{\text{local}}, \end{aligned}$$

where $\mathbf{x}_j = (x_j, y_j, z_j)^\top$ is the position of the $j$th magnet and $\mathbf{m}_j = m_j(\sin\theta_j \cos\phi_j, \sin\theta_j \sin\phi_j, \cos\theta_j)^\top$ is its magnetization vector, where $\theta_j$ and $\phi_j$ are the pitch and yaw of the magnet, respectively.

We wished to know what the accuracy of the magnet tracking would be if all the magnetometers were used at once to track all the magnets at once (global tracking), as opposed to just tracking each magnet pair using one magnetometer array with only the local orientation corrections applied (independent tracking). To compare the accuracy of global tracking versus independent tracking, we used our residual limb model. We first created ground truth measurements by individually tracking each magnet pair with no neighboring magnets present, calculating the distance between the magnets in each pair. We then simultaneously implanted all magnet pairs into the residual limb model and compared the distance errors from global tracking against the distance errors from simultaneous independent tracking of each magnet pair. We first performed this with a static test, then with a dynamic test simulating residual limb pistoning by physically cyclically translating the residual limb model multiple centimeters vertically (along its x-axis).

The Kabsch algorithm successfully aligned the magnetic fields of the four magnetometer arrays. The alignment of the fields can be observed in the before and after images, with both the three-dimensional and time-series views showing the alignment (Fig. S6). The three non-reference magnetometer arrays were found to have orientation magnitudes of 72.27, 177.66, and 140.35 degrees relative to the reference array (Fig. S6). The local orientations were found to be repeatable with sub-degree accuracy (Fig. S7). Specifically, the median RMS angular errors for the four arrays were 0.14, 0.18, 0.13, and 0.12 degrees, respectively, while the median local orientation magnitudes were 0.41, 0.51, 0.52, and 0.47 degrees, respectively, with the median error in all cases being less than the median magnitude. The orientation calibration of the full dataset completed in less than one second (running as unoptimized Python code). The position calibration was found to have a repeatability of better than 0.4 mm (position standard deviation across repeated tests) in all cases, with clear visual alignment of the tracked paths of the reference magnet (Fig. S8). Not accounting for the time to track the reference magnet (which can be performed in real-time when tracked via a real-time system), the position calibration completed in less than two seconds (also running as unoptimized Python code). The static tracking test demonstrated a submillimeter tracking accuracy for the global tracking after magnetometer poses calibration (Fig. S9), in comparison with ground truth. Specifically, global tracking errors in the distance-between-magnets signal were -0.17, 0.20, -0.62, and 0.01 mm, while the independent tracking errors were substantially higher: 0.49, -2.01, -6.42, and -1.96 mm, respectively. The dynamic tracking test demonstrated even more dramatic differences between the global and independent tracking (Fig. S10), with the global tracking errors staying substantially more stable than their diverging independent tracking counterparts.

**Supplementary Note 2: Information filter description and implementation**

The strategy applied to the state dynamics is to model the magnet movement as a random process. The magnet dynamics are driven by a second-order Markov process which inputs process noise into the system. This second-order Markov process drives the noise in the derivative (velocity) states; when propagated through the state transition this produces a correlated random-walk in the primary position and orientation states, accurately capturing the stochastic motion of each magnet.

When tracking both the position and orientation (dipole moment) of each magnet, the state vector is extended to incorporate complementary states derived from the second-order Markov model and disturbance states representing the estimated uniform geomagnetic field.

For $M$ magnets indexed by $j = 1, \ldots, M$, each magnet's primary state vector $\mathbf{x}_j$ is:

$$\mathbf{x}_j^\mathsf{T} = [x_j \quad y_j \quad z_j \quad \theta_j \quad \phi_j]$$

The vector $\mathbf{x}_j$ contains the *primary* states of the $j$-th magnet, namely its position coordinates $(x_j,\ y_j,\ z_j)$ and the two angles $(\theta_j,\ \phi_j)$ that specify the direction of its magnetic moment dipole vector.

The auxiliary vector $\tilde{\mathbf{x}}$ holds the *complementary* states introduced by the second-order Markov process. These states are the first derivatives (velocities) of the primary variables and where the process noise is injected into, thereby propagating uncertainty from the velocity level into the position and orientation components. Each primary state, including the geomagnetic disturbance, has its own complementary state.

The complementary states derived from the second-order Markov model serve to capture the temporal dynamics and uncertainties associated with the primary states. By modeling the system as a second-order process, the filter accounts for both the current state and its rate of change, enabling more accurate predictions and smoother state estimates. The second-order Markov model facilitates the propagation of process noise through the system, inducing a "random walk" behavior in the primary states. This stochastic behavior enables the filter to adapt to gradual changes and maintain flexibility in tracking the magnet's motion. Incorporating complementary states allows for a more structured covariance matrix, capturing the correlations between positions, orientations, and their derivatives. This leads to improved estimation accuracy and robustness against uncertainties. In addition, this is also a unique way of being able to compute velocity as well rather than methods susceptible to noise, such as a finite-difference derivative.

The vector $\mathbf{x}_{\text{geo}}$ represents the *disturbance* states that are the geomagnetic field and any other slowly varying environmental magnetic biases. Including these terms in the state vector enables the filter to estimate and compensate external disturbances concurrently with the primary and complementary states, improving the accuracy of magnetic dipole state estimates.

Each sensor board is assigned the three states that represent the local, approximately uniform geomagnetic bias measured by that board. Assume $B$ boards, indexed by $b \in \{1, \dots, B\}$, each consisting of an array of magnetic field sensors. Depending on the number of boards, the disturbance vector becomes

$$\mathbf{x}_{\text{geo}}^{\mathsf{T}} = [G_{1x} \quad G_{1y} \quad G_{1z} \quad \cdots \quad G_{Bx} \quad G_{By} \quad G_{Bz}], \qquad G_{bx}, G_{by}, G_{bz} \in \mathbb{R},$$

Let $\mathbf{g}_b = [\,G_{bx},\ G_{by},\ G_{bz}\,]^{\mathsf{T}} \in \mathbb{R}^3$ denote the local, approximately uniform geomagnetic field measured by sensor-board $b \in \{1, \dots, B\}$. Stacking the $B$ board vectors give

$$\mathbf{x}_{\text{geo}}^{\mathsf{T}} = [\mathbf{g}_1^{\mathsf{T}} \quad \cdots \quad \mathbf{g}_B^{\mathsf{T}}] \in \mathbb{R}^{3B}.$$

Disturbance states model the uniform geomagnetic field and any other external magnetic disturbances that affect sensor measurements. The inclusion is critical as by estimating and compensating for external magnetic fields, the filter ensures that the primary state estimates are not biased or distorted by environmental factors. Accounting for this leads to more reliable sensor measurements, as the filter can subtract the estimated disturbances from the raw measurements, isolating the magnetic field contributions from the tracked magnets. Modeling disturbances as part of the state vector allows the filter to adapt to varying environmental conditions, maintaining consistent tracking performance across varying orientations relative to the geomagnetic field. The disturbance states also have their respective complementary states $\tilde{\mathbf{x}}_{\text{geo}}^{\mathsf{T}}$.

The comprehensive state vector $\mathbf{x}$ combines all the previously defined components:

so that the full state vector is

$$\mathbf{x}^{\mathsf{T}} = \left[\mathbf{x}_1^{\mathsf{T}} \ \dots \ \mathbf{x}_M^{\mathsf{T}} \ \tilde{\mathbf{x}}^{\mathsf{T}} \ \mathbf{x}_{\text{geo}}^{\mathsf{T}} \ \tilde{\mathbf{x}}_{\text{geo}}^{\mathsf{T}}\right]$$

During filtering, these components absorb slow variations of the geomagnetic field and other quasi-static magnetic biases local to each board, thereby preventing such disturbances from corrupting the magnet state estimates, and allowing the filter to predict magnet estimates at distances away from the board that would cause the strength generated by the dipoles to be less than the geomagnetic field.

The process model governs the evolution of the state vector over time, incorporating both the dynamics of the magnetic dipole and process noise. This serves as the state propagation layer. The discrete-time state transition is modeled as:

$$\mathbf{x}_k = \mathbf{F}\mathbf{x}_{k-1} + \mathbf{G}\mathbf{w}_{k-1}$$

where **F** is the state transition matrix, **G** is the input matrix, and $\mathbf{w}_{k-1} \sim \mathcal{N}(\mathbf{0}, \mathbf{Q})$ is the process noise, with state covariance matrix **Q**.

The state transition matrix **F** is constructed based on the time constant $\tau$ and the timestep $dt$:

$$\mathbf{F} = e^{\mathbf{A} \cdot dt}$$

where **A** is the continuous-time state transition matrix defined as:

$$\mathbf{A} = \begin{bmatrix} -\frac{1}{\tau}\mathbf{I} & \frac{1}{\tau}\mathbf{I} \\ \mathbf{0} & -\frac{1}{\tau}\mathbf{I} \end{bmatrix}$$

**A** has size $N \times N$, where $N = 2(5M + dB)$, $d = 3$. The factor $5M$ counts *primary* $(x, y, z, \theta, \phi)$ states for the $M$ magnets, $dB = 3B$ accounts for the geomagnetic vectors $\mathbf{g}_b$, and the leading factor 2 adds the corresponding complementary (velocity) states introduced by the second-order Markov model. Hence **A**, **F**, and **Q** are all $N \times N$ matrices that are always the size of the total states.

The continuous-time process is equivalent to two identical first-order low-pass filters cascaded in series (a critically damped double integrator), each with time constant $\tau$. After discretization with sampling period $dt$ the corresponding poles are located at $e^{-dt/\tau}$. Increasing $\tau$ moves the poles closer to unity, so the filter averages a longer window of time-correlated measurements: the process noise injected into the velocity complementary state is attenuated twice before it reaches the position state, reducing the estimator's variance but narrowing the bandwidth of response. Conversely, decreasing $\tau$ raises the effective cut-off frequency, yielding faster response to rapid motion at the cost of higher estimation variance. Selecting $\tau$ to match the dominant time scale of the muscle movement (or other application) therefore provides a principled trade-off between responsiveness and precision. This intuition drives the state propagation model.

The process noise covariance matrix **Q** accounts for uncertainties in the state dynamics and is composed of process noise terms for the primary states: position, orientation, and geomagnetic disturbance components. All entries associated with the complementary (velocity) states are set to zero.

The discrete-time noise–input matrix **G** is part of the second–order Markov model that injects white process noise $\mathbf{w}_{k-1}$ only into the complementary *velocity* layer, so that for every scalar pair $(x, \tilde{x})$

$$\begin{bmatrix} x_k \\ \tilde{x}_k \end{bmatrix} = \begin{bmatrix} -1/\tau & 1/\tau \\ 0 & -1/\tau \end{bmatrix} \begin{bmatrix} x_{k-1} \\ \tilde{x}_{k-1} \end{bmatrix} + \begin{bmatrix} 0 \\ 1/\tau \end{bmatrix} \mathbf{w}_{k-1}, \qquad \mathbf{w}_{k-1} \sim \mathcal{N}(\mathbf{0}, \mathbf{Q})$$

In the discretized second-order Markov model the noise–input matrix **G** has a very specific structure: every column that corresponds to a *primary* state is zero, whereas the rows associated with the complementary (velocity) states contain the non-zero entries $1 - e^{-dt/\tau}$ and $\tau\left(1 - e^{-dt/\tau}\right)$ derived from the continuous-to-discrete conversion. Thus the white-noise sequence $\mathbf{w}_k$ is injected *only* into the velocity layer; at the next prediction step the state-transition matrix $\mathbf{F}$ integrates that perturbation forward into the position–orientation layer. This arrangement recreates the intended second-order, correlated random walk that suppresses high-frequency jitter in the primary states while still allowing quick response driven by the process noise.

The measurement model relates the state vector to the sensor measurements, which are the magnetic field vectors observed by each sensor. The measurement at timestep $k$ is given by:

$$\mathbf{z}_k = \mathbf{h}(\mathbf{x}_k) + \mathbf{v}_k$$

where $\mathbf{h}(\mathbf{x}_k)$ is the nonlinear measurement function and $\mathbf{v}_k \sim \mathcal{N}(\mathbf{0}, \mathbf{R})$ is the measurement noise, with noise covariance matrix $\mathbf{R}$ of size $I \times 3$ for $I$ magnetic field sensors, each providing 3-axis measurements.

The magnetic field generated by a dipole at a sensor location is given by the fundamental dipole field equation:

$$\mathbf{B}_j^i = \frac{\mu_0}{4\pi}\left(\frac{3\mathbf{r}_{ij}(\mathbf{m}_j \cdot \mathbf{r}_{ij})}{|\mathbf{r}_{ij}|^5} - \frac{\mathbf{m}_j}{|\mathbf{r}_{ij}|^3}\right)$$

where $\mu_0$ is the permeability of free space, $\mathbf{r}_{ij} = [s_x^i - x_j,\ s_y^i - y_j,\ s_z^i - z_j]^\mathsf{T}$ is the displacement vector from dipole $j$ to sensor $i$, and $\mathbf{m}_j = [m_{xj},\ m_{yj},\ m_{zj}]^\mathsf{T}$ is the dipole-moment vector of magnet $j$.

The corresponding unit vector $\hat{\mathbf{m}}_j$ is obtained by the standard spherical-to-Cartesian mapping:

$$\hat{\mathbf{m}}_j = \begin{bmatrix} \sin\theta_j \cos\phi_j \\ \sin\theta_j \sin\phi_j \\ \cos\theta_j \end{bmatrix} = \begin{bmatrix} \hat{m}_{xj} \\ \hat{m}_{yj} \\ \hat{m}_{zj} \end{bmatrix}$$

These equation calculates the estimated magnetic field vector at sensor $i$ given the position and magnetic moment vector of dipole $j$, based on the distance vector between the magnet and sensor locations in the defined reference frame. Throughout this work the magnitude of every magnet's dipole moment is treated as a *known* scalar rather than an estimated state. The permeability of free space $\mu_0$ and any other scalar terms are implicitly included into this lumped value. In all field equations we therefore write: $\mathbf{m}_j = m_0\, \hat{\mathbf{m}}_j$ where $\hat{\mathbf{m}}_j$ is the unit vector encoded by the orientation states $(\theta_j, \phi_j)$. Process noise is not applied to $m_0$, reflecting the assumption that the magnetization of each bead remains constant. The total magnetic field at sensor $i$ is the sum of contributions from all dipoles and any disturbance:

$$\mathbf{z}^i = \sum_{j=1}^{M} \mathbf{B}_j^i + \mathbf{G}^i + \mathbf{v}^i$$

where $\mathbf{G}^i$ represents the external disturbance (such as the geomagnetic field) at sensor $i$. Accurate computation of the Jacobian matrix $\mathbf{H}_k$ is critical for the tracking performance. The Jacobian is calculated due to the nonlinearities in the measurement model $\mathbf{h}(\mathbf{x}_k)$ as a way to linearize the function. The nominal operation point that the Jacobian is linearized about is the *a priori* (predicted) state:

$$\mathbf{x}_{k|k-1} = \mathbf{F}\,\mathbf{x}_{k-1|k-1}$$

The resulting Jacobian is

$$\mathbf{H}_k = \frac{\partial \mathbf{h}}{\partial \mathbf{x}}\bigg|_{\mathbf{x}=\mathbf{x}_{k|k-1}} \in \mathbb{R}^{m\times n}$$

where $n$ and $m$ are the dimensions of the state and measurement vectors, respectively. Accurate, analytical evaluation of $\mathbf{H}_k$ is essential: it supplies the first-order sensitivity of the measurement to the state and determines the information gain used to map the measurement residual back into state space. Any error in $\mathbf{H}_k$ therefore propagates directly into the state update and degrades tracking accuracy. Therefore, we analytically compute the Jacobian rather than relying on numerical methods such as finite difference which reduce accuracy and increase latency.

Each dipole contributes the five *primary* states $(x_j, y_j, z_j, \theta_j, \phi_j)$ and the corresponding five *complementary* (velocity) states. With $I$ sensors, $M$ dipoles and $B$ sensor boards the measurement Jacobian has

$$\mathbf{H}_k \in \mathbb{R}^{3I\times(5M+5M+3B)}$$

Using the sensor index $i = 1, \ldots, I$ and dipole index $j = 1, \ldots, M$ we write

$$\mathbf{H}_k = \begin{bmatrix} \mathbf{H}_{11} & \ldots & \mathbf{H}_{1M} & \mathbf{0} & \ldots & \mathbf{0} & \mathbf{G}_1 \\ \mathbf{H}_{21} & \ldots & \mathbf{H}_{2M} & \mathbf{0} & \ldots & \mathbf{0} & \mathbf{G}_2 \\ \vdots & \ddots & \vdots & \vdots & \ddots & \vdots & \vdots \\ \mathbf{H}_{I1} & \ldots & \mathbf{H}_{IM} & \mathbf{0} & \ldots & \mathbf{0} & \mathbf{G}_I \end{bmatrix}$$

where $\mathbf{H}_{ij} \in \mathbb{R}^{3\times5}$ carries the derivatives of the field measured by sensor $i$ with respect to the five primary states of dipole $j$; the $3I \times 5M$ zero block corresponds to the complementary states, for which $\partial\mathbf{h}/\partial\tilde{\mathbf{x}} = \mathbf{0}$; $\mathbf{G}_i \in \mathbb{R}^{3\times3B}$ inserts the $3 \times 3$ identity in the three columns associated with the board that contains sensor $i$ (geomagnetic disturbance states) and zeros elsewhere. The assumption is that the disturbance field is seen uniformly across all the sensors in the same way, resulting in the Jacobian being simplified to an identity matrix. For a particular sensor–dipole pair the $3 \times 5$ block is

$$\mathbf{H}_{ij} = \begin{bmatrix} \frac{\partial \mathbf{B}_j^i}{\partial x_j} & \frac{\partial \mathbf{B}_j^i}{\partial y_j} & \frac{\partial \mathbf{B}_j^i}{\partial z_j} & \frac{\partial \mathbf{B}_j^i}{\partial \theta_j} & \frac{\partial \mathbf{B}_j^i}{\partial \phi_j} \end{bmatrix}$$

Let $\mathbf{r}_{ij} = \mathbf{s}_i - \mathbf{p}_j$, $r = \| \mathbf{r}_{ij} \|$, and write the fixed-magnitude dipole moment as $\mathbf{m}_j = m_0\hat{\mathbf{m}}_j$, where $\hat{\mathbf{m}}_j$ is the unit vector encoded by the orientation states $(\theta_j, \phi_j)$.

The magnetic-field measurement function $\mathbf{h}(\mathbf{x})$ depends only on the current positions, orientation angles, and disturbance field estimation; it is independent of the velocity (complementary) states introduced for the second-order Gauss–Markov process. Therefore $\partial\mathbf{h}/\partial\tilde{\mathbf{x}} = \mathbf{0}$, and the corresponding columns in $\mathbf{H}_k$ are identically zero. The magnetic-field measurement $\mathbf{h}(\mathbf{x})$ depends only on the *instantaneous* positions, orientations, and board-level geomagnetic biases; it contains no explicit dependence on the velocity ("complementary") states that were introduced solely to model the process dynamics. Consequently, the partial derivatives with respect to the complementary states are zero: $\frac{\partial \mathbf{h}}{\partial \tilde{\mathbf{x}}} = \mathbf{0}$, and the corresponding columns in $\mathbf{H}_k$ are identically zero. Physically this is consistent with the fact that a magnetic field sensor responds to the magnetic field produced by the magnets' present configuration, not to how fast that field is changing (which would be a derivative of the flux).

**Implementation**

The extended information filter (INFO) operates with the information matrix $\mathbf{Y}$ and the information vector $\mathbf{y}$, which are the inverse of the covariance matrix $\mathbf{P}$ and the product of $\mathbf{P}^{-1}$ with the state mean $\hat{\mathbf{x}}$, respectively. Large diagonal values within the covariance matrix intuitively imply a lack of precision or a larger degree of uncertainty regarding those state estimate components. Conversely, large diagonal elements imply a high degree of precision, or a smaller degree of uncertainty, of those state estimate components. Think of the covariance $\mathbf{P}$ as describing state uncertainty, the goal is to minimize the covariance matrix. When viewing the problem from the information lens, the goal is to maximize information gain $\mathbf{Y}$.

First, to initialize the state vector $\mathbf{x}_0$, the covariance matrix $\mathbf{P}_0$, and compute the initial information matrix and vector $\mathbf{Y}_0$ and $\mathbf{y}_0$, the system can be driven into steady state by

$$\mathbf{P}_{k+1} = \mathbf{F}\,\mathbf{P}_k\,\mathbf{F}^\top + \mathbf{G}\,\mathbf{Q}\,\mathbf{G}^\top, \qquad k = 0,1,\ldots$$

starting from $\mathbf{P}_0 = \mathbf{I}$ and continuing until $\mathbf{P}$ has reached steady-state (large number of $k$ iterations). And, for the full state vector of dimension $N = 2(5M + 3B)$,

$$\mathbf{P}_0 = \mathbf{P}^\star \in \mathbb{R}^{N\times N}$$

Finally, we convert the steady-state covariance to information form:

$$\boxed{\mathbf{Y}_0 = \mathbf{P}_0^{-1}, \qquad \mathbf{y}_0 = \mathbf{Y}_0\,\mathbf{x}_0}$$

so that the filter starts with the maximum *a-priori* information consistent with the assumed process model and noise.

In the prediction step, the filter propagates the state and information matrices forward based on the previously described process model. The state prediction incorporates the state transition matrix $\mathbf{F}$, the process noise covariance $\mathbf{Q}$ and process input $\mathbf{G}$.

$$\begin{aligned}
\mathbf{x}_{k|k-1} &= \mathbf{F}\mathbf{x}_{k-1|k-1} \\
\mathbf{P}_{k|k-1} &= \mathbf{F}\mathbf{P}_{k-1|k-1}\mathbf{F}^\top + \mathbf{G}\mathbf{Q}\mathbf{G}^\top \\
\mathbf{Y}_{k|k-1} &= \mathbf{P}_{k|k-1}^{-1} \\
\mathbf{y}_{k|k-1} &= \mathbf{Y}_{k|k-1}\mathbf{x}_{k|k-1}
\end{aligned}$$

The inverse of the predicted covariance matrix $\mathbf{Y}_{k|k-1}$ is computed by propagating the previous covariance through the state transition matrix $\mathbf{F}$ and adding the process noise covariance $\mathbf{Q}$ through process input matrix $\mathbf{G}$. The predicted information vector $\mathbf{y}_{k|k-1}$ is obtained by multiplying the predicted information matrix $\mathbf{Y}_{k|k-1}$ with the propagated state vector $\mathbf{F}\mathbf{x}_{k-1|k-1}$. This step prepares the information vector for incorporation of new measurements. Defining the measurement residual $\boldsymbol{\nu}_k = \mathbf{z}_k - \mathbf{h}(\hat{\mathbf{x}}_{k|k-1})$ and the *augmented innovation*

$$\tilde{\mathbf{z}}_k = \boldsymbol{\nu}_k + \mathbf{H}_k\,\hat{\mathbf{x}}_{k|k-1}, \qquad \mathbf{H}_k = \left.\frac{\partial \mathbf{h}}{\partial \mathbf{x}}\right|_{\hat{\mathbf{x}}_{k|k-1}}$$

The information-domain update is then

$$\mathbf{S}_k = \mathbf{H}_k^{\mathsf{T}} \mathbf{R}^{-1} \mathbf{H}_k,$$
$$\mathbf{Y}_k = \mathbf{Y}_{k|k-1} + \mathbf{S}_k,$$
$$\mathbf{y}_k = \mathbf{y}_{k|k-1} + \mathbf{H}_k^{\mathsf{T}} \mathbf{R}^{-1} \tilde{\mathbf{z}}_k,$$

Here $\mathbf{S}_k$ is the innovation information contributed by the new measurement, while $\mathbf{H}_k^{\mathsf{T}} \mathbf{R}^{-1} \tilde{\mathbf{z}}_k$ is the information *increment* derived from the residual. When $\mathbf{h}(\mathbf{x})$ is linear the term $\mathbf{H}_k \hat{\mathbf{x}}_{k|k-1}$ cancels, recovering the standard linear information filter equations. This additional term is an artifact of the first-order Taylor series used to convert the nonlinear measurement function into the equivalent *linear* form required by the information space matrix operations.

This concise formulation shows that all covariance and residual effects are accumulated in information space; the updated mean and covariance are recovered only once per timestep *dt* through the inversion of $\mathbf{Y}_k$. The state estimate update step finalizes the incorporation of the new measurements by computing the updated state vector and covariance matrix from the information matrix and information vector.

$$\mathbf{x}_{k|k} = \mathbf{Y}_k^{-1} \mathbf{y}_k$$
$$\mathbf{P}_{k|k} = \mathbf{Y}_k^{-1}$$

The updated state estimate $\mathbf{x}_{k|k}$ is obtained by multiplying the inverse of the updated information matrix $\mathbf{Y}_k^{-1}$ with the updated information vector $\mathbf{y}_k$. This operation yields the most probable state estimate given the prior predictions and the new measurements. The updated covariance matrix $\mathbf{P}_{k|k}$ is simply the inverse of the updated information matrix $\mathbf{Y}_k^{-1}$. This matrix represents the uncertainty associated with the updated state estimate. The process then loops again for the next state estimate.

**Supplementary Note 3: Biophysical model for kinematic reconstruction**

To estimate joint kinematics from muscle activation signals (sEMG/iEMG), we implemented a learnable, second-order forward-dynamics model. The model describes a single-degree-of-freedom joint controlled by a pair of muscles. The core of the system is modeled as a damped harmonic oscillator, whose dynamics are governed by the following second-order linear differential equation:

$$\begin{bmatrix} \dot{\theta} \\ \ddot{\theta} \end{bmatrix} = \begin{bmatrix} 0 & 1 \\ -\frac{K}{I} & -\frac{B}{I} \end{bmatrix} \begin{bmatrix} \theta \\ \dot{\theta} \end{bmatrix} + \begin{bmatrix} 0 \\ \frac{1}{I} \end{bmatrix} \tau_m$$

In this equation, $\theta$ represents the joint angle, $I$ is the effective moment of inertia of the joint, $K$ is the total stiffness of the joint, $B$ is the damping coefficient, and $\tau_m(t)$ is the net torque generated by the muscle pair that drives the joint toward an equilibrium angle. The model's parameters are dynamically modulated by the muscle activations: $a_1(t)$ and $a_2(t)$, which are derived from the processed EMG envelopes from the muscle pair.

The total stiffness is the sum of the individual muscle stiffnesses. Each muscle's stiffness is a linear function of its activation:

$$K_{joint}(t) = (K_{0,1} + K_{1,1} \cdot a_1(t)) + (K_{0,2} + K_{1,2} \cdot a_2(t))$$

Here, $K_0$ represents the passive stiffness of the muscle, and $K_1$ is the gain that determines how much stiffness increases with activation. The subscripts 1 and 2 denote the two antagonistic muscles in the pair. The model assumes the joint is critically damped to produce smooth, non-oscillatory motion. The damping coefficient is therefore dependent on the dynamic stiffness and inertia:

$$b(t) = 2\sqrt{K_{joint}(t) \cdot I}$$

The net torque is determined by the balance of forces from the antagonistic muscles. Each muscle has an activation-dependent equilibrium angle, $L(t)$. The product of a muscle's stiffness and its equilibrium angle gives its contribution to the torque.

$$\tau(t) = [K_1(t) \cdot L_1(t)] - [K_2(t) \cdot L_2(t)]$$

where the equilibrium angle for each muscle, $L(t)$, is also a linear function of its activation:

$$L_i(t) = L_{0,i} + L_{1,i} \cdot a_i(t) \quad \text{for } i \in \{1,2\}$$

$$K_i(t) = K_{0,i} + K_{1,i} \cdot a_i(t) \quad \text{for } i \in \{1,2\}$$

Here, $L_0$ is the passive equilibrium angle and $L_1$ is the activation-dependent change in that angle. The parameters of this model were optimized to fit the experimental data. The set of learnable parameters includes muscle parameters and joint parameters.

For each of the two antagonistic muscles, four parameters were learned: passive stiffness ($K_0$), active stiffness gain ($K_1$), passive equilibrium angle ($L_0$), and gain in active equilibrium angle ($L_1$).

The joint parameters included both moment of inertia (I) and an additional scaling parameter representing the moment arm that transforms the linear space of the muscle pair to the rotary space of the joint.

For each trial, the model's parameters were trained to minimize the error between its predicted kinematic trajectory and the reference trajectory recorded by the goniometer. The full 30-second window of each trial was used to train the model. The optimization objective was to minimize the mean squared error between the model's predicted joint angle and the ground-truth goniometer angle. The optimizer had a fixed learning rate of 0.001, and the model was trained for 500 epochs which demonstrated loss convergence for each of the trials

**Supplementary Note 4: Neuromuscular controller**

Activation signals from EMG served as inputs to a controller design similar to Song *et al.*[1] that simulates the real-time dynamics of two AMI muscle pairs controlling independent DOFs for both position and impedance control. Intended target position was computed by the difference of the weighted muscle pair activities:

$$[(K_{0,\text{agonist}} + K_{1,\text{agonist}} \cdot a_{\text{agonist}}) \cdot (x_{\text{eq,agonist}} + c_{\text{agonist}} \cdot a_{\text{agonist}}) - (K_{0,\text{antagonist}} + K_{1,\text{antagonist}} \cdot a_{\text{antagonist}}) \cdot (x_{\text{eq,antagonist}} + c_{\text{antagonist}} \cdot a_{\text{antagonist}})],$$

where the stiffness component combined a baseline stiffness value ($K_0$) with an activation-scaled stiffness increment ($K_1 \cdot a$), and a represents the normalized EMG activation ranging from 0 to 1. The equilibrium point incorporates both the resting point at zero activation ($x_{eq}$) and an activation-dependent linear scale factor in changing the equilibrium point ($c \cdot a$). The impedance setpoint was determined by the combined output of the stiffness contribution:

$$(K_{0,\text{agonist}} + K_{1,\text{agonist}} \cdot a_{\text{agonist}}) + (K_{0,\text{antagonist}} + K_{1,\text{antagonist}} \cdot a_{\text{antagonist}})$$

For the MuSE controller, each muscle was assigned a defined resting position, and cursor control was achieved through changes from these resting muscle states to a defined maximum deflection. For ankle joint control, resting positions were defined as the resting muscle length (distance between the two beads). Muscle shortening was quantified by measuring displacement from these rest positions, where only muscle shortening (decreased distance) contributed to deflection. The deflection was normalized by dividing a maximum calibrated deflection and scaled by muscle-specific gains. The resulting intended target position was computed by taking the difference between the normalized, scaled tibialis anterior and gastrocnemius deflections. Similarly, for subtalar joint control, individual magnets associated with inversion and eversion movements each had calibrated resting positions along the chosen tracking axes, with the axis selected based on the highest signal-to-noise ratio for the intended muscle movement. Movement away from these rest positions generated control signals by calculating the difference between the measured and resting positions, normalized by the maximum deflection and scaled by muscle-specific gains. The subtalar target position was then computed as the difference between the tibialis posterior and peroneus longus components.

**Supplementary Note 5: Surgical details and implementation**

The surgical implantation of the magnetic beads and e-OPRA systems was split across two separate stage procedures that were spaced 3-6 months apart. The first stage encompassed: (i) soft tissue muscle constructs; (ii) osteotomy; and (iii) implantation of titanium BioHelix fixture, intramuscular electrodes and magnetic beads. Initially, muscles were dissected to create two AMI pairs: lateral gastrocnemius (GAS) and tibialis anterior (TA) for prosthetic ankle joint control, and tibialis posterior (TP) and peroneus longus (PL) for subtalar joint control. After AMI creation and osteotomy, subsequent interface implantation included both permanent implanted electrodes and magnetic beads. Two magnets were implanted into each ankle AMI muscle along its longitudinal axis. Each muscle's bead pair was used to wirelessly track that muscle's length and speed, while 8 bipolar implanted electrode pairs captured isolated muscle activation of each AMI muscle construct. For all implanted magnets, the target separation distance for any magnet-to-magnet pair was 4 cm for all muscle contractile states to prevent bead migration within the first six weeks. Due to this limitation, only one magnet could be implanted into each of the subtalar muscles, TP and PL, to prevent exceeding the required separation distance within the residual limb. Magnets were implanted using one of two approaches: either through a small incision to create a muscle pocket for bead placement, or by direct insertion into the muscle tissue. Bead pairs were implanted within the muscle belly of each ankle AMI muscle and aligned along individual muscle fibers to maximize movement resolution. Implantation was performed using either a bead inserter (Halifax Biomedical) or nonmagnetic surgical tweezers. Both subjects 1 and 2 received a total of 6 magnets; two for each ankle AMI muscle and one for each subtalar AMI muscle. As subject 3 was an amputation revision procedure, only 2 magnets were placed in each ankle AMI muscle for a total of 4 magnets, as subsequent scar tissue prevented surgical access to the deeper subtalar muscles.

Following an initial recovery from the first stage, subject 1 and 2 received the second stage which involved placing the eAbutment into the BioHelix fixture, allowing both (i) accessibility to the implanted electrode connections through a 16-pin connector and (ii) mechanical connection of an external prosthesis to the osseointegrated implant. No complications, adverse events or subject discomfort were related to the magnetic beads. No bead migration occurred in all 3 subjects. Subject 2 had to receive a tightening of their fixture and abutment, while subject 1 has had no complications with the e-OPRA system. Subject 3 had to receive a fixture replacement, thus their implanted electrodes were not accessible for this study.

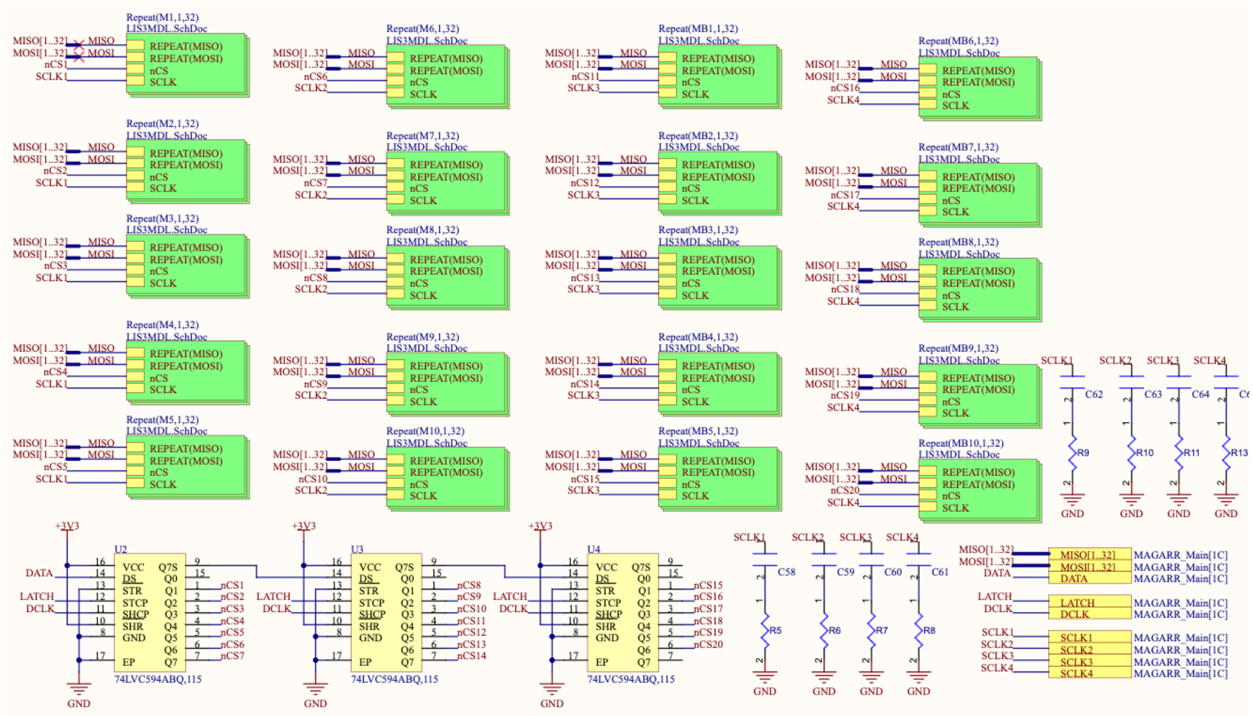


**Figure S1. Schematic of sensor logic**

Sensor array containing 640 LIS3MDL magnetometers distributed across 32 SPI channels operating at 10 MHz clock frequency for high-density magnetic field sampling. Cascaded shift register that provides distribution of the chip select signals across the sensors. Each repeated block contains 32 sensors that run on SPI data line, while the clock signal is evenly distributed across 5 of the blocks.

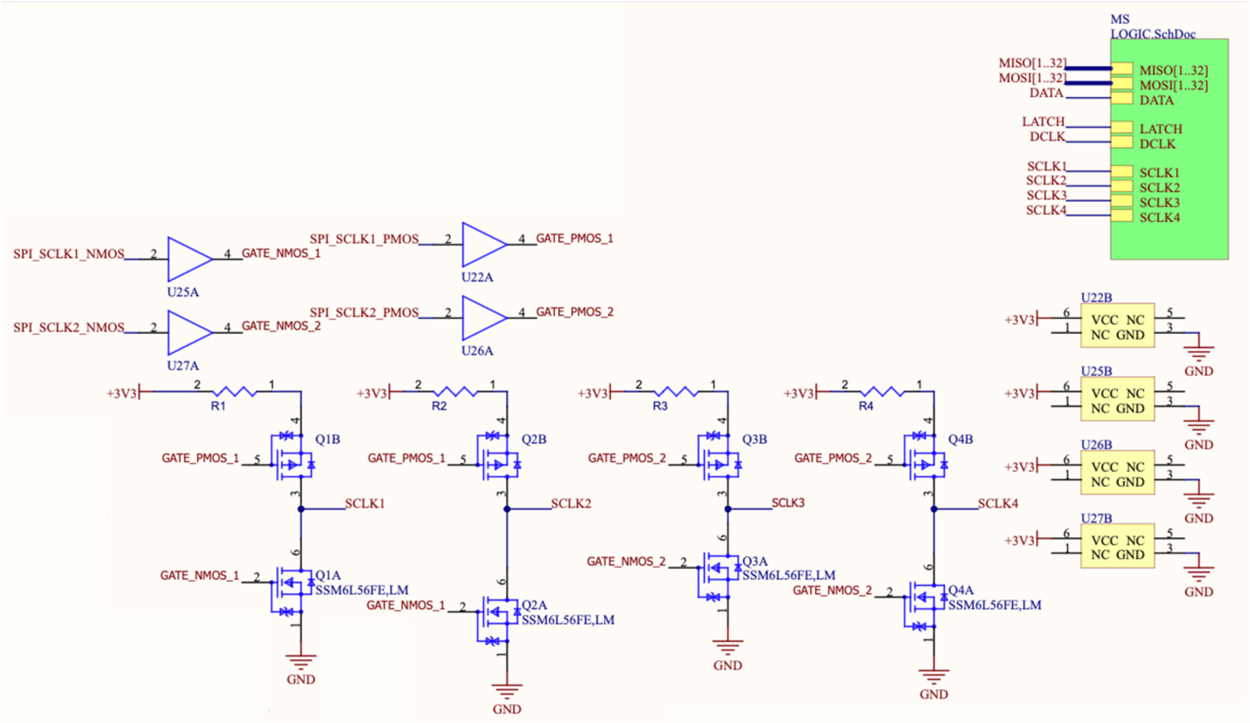


**Figure S2. Schematic of clock driver circuit.** Four identical clock distribution channels driving all the magnetometers. Each SPI clock signal passes through a voltage level translator to step up from 1.8V FPGA logic to 3.3V sensor levels, then drives a complementary push-pull MOSFET stage. The push-pull driver consists of MOSFETs configured to actively drive both rising and falling edges, providing high current capability for the cumulative capacitive load of many sensors per clock line.

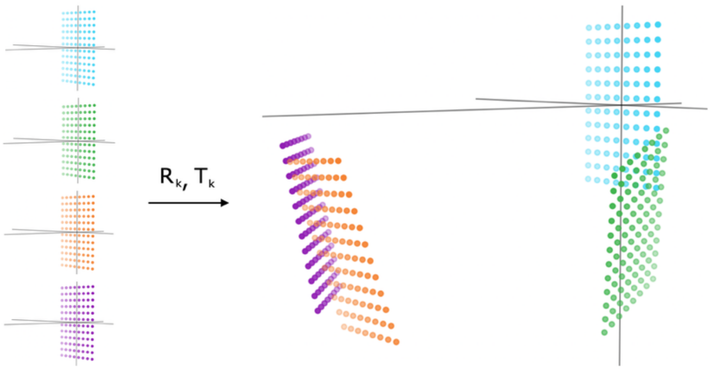


**Figure S3. Magnetometer calibration**

Before calibration, the magnetometer arrays each have their own coordinate frame. The magnetometer array poses calibration determines the relative orientation and position of each magnetometer array so that they can be transformed into and share a global frame.

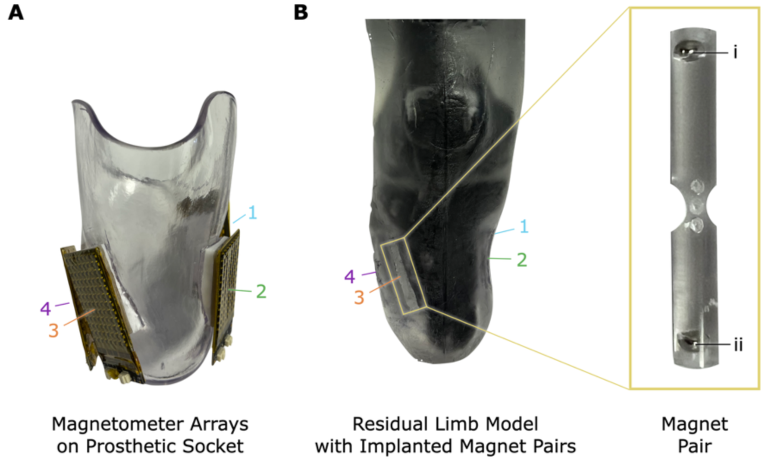


**Figure S4. Benchtop setup to validate magnetometer calibration**

(a) To test the repeatability of the proposed calibration, we rigidly affixed four 96-magnetometer arrays to a prosthetic socket. (b) To test the accuracy of magnet tracking with and without the proposed calibration, we created a residual limb model from the MRI scan of a patient and implanted four pairs of magnets into muscle locations that would be used to control a two-degree of freedom prosthesis. Numbered magnet pairs in the right image correspond to numbered arrays in the left image, and two magnets in a representative acrylic fixture are shown with labels i and ii.

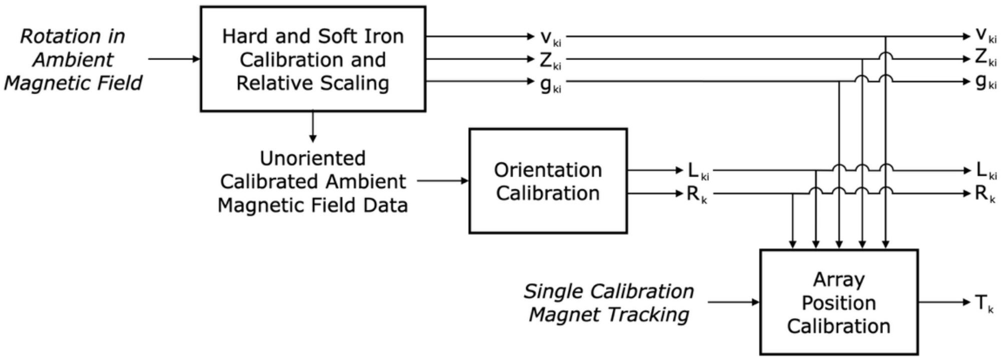


**Figure S5. Calibration process**

This block diagram shows the process for implementing the proposed calibration for the *i*th magnetometer and *k*th array. First, the rigidly-affixed magnetometer arrays are rotated together in a spatially-uniform ambient magnetic field to perform the traditional hard and soft iron calibration and to scale the magnetometers relative to one another. The parameters from this first calibration step are applied to the initial dataset, and a second calibration step is performed to determine the orientations of the magnetometers, aligning all magnetometers relative to one another. Finally, a second calibration dataset is collected using a calibration magnet, and the calibrated (**v** and **Z**), scaled (*g*), and oriented (**L** and **R**) magnetic field measurements from this final calibration dataset are used to determine the positions (**p**)of the magnetometer arrays.

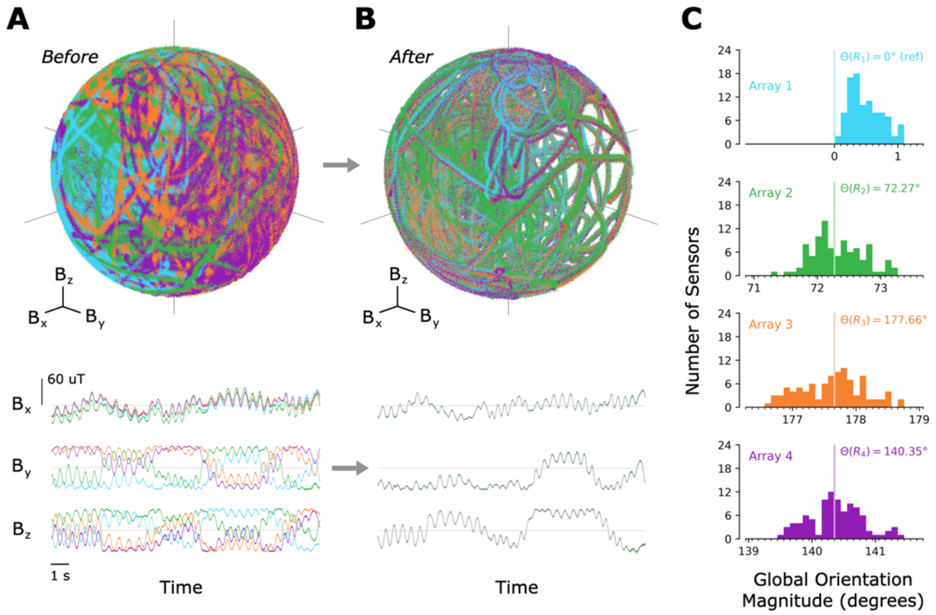


**Figure S6. Magnetometer orientation calibration of four magnetometer arrays**

(a) Upon completing the hard- and soft-iron calibrations and relative scaling from the ambient magnetic field rotation dataset, the data from the four magnetometer arrays can be seen as a set of concentric spheres, as seen in the three-dimensional plot on top, but without the fields aligned with one another, as can be seen in a representative section of the time-series plots on bottom. Data from all 96 magnetometers of each array is plotted here, though only four curves seem to appear due to the alignment of the magnetometers on each array. (Note also that the similarity in data between the four arrays is due to the magnetometer arrays being similarly oriented in the x-direction.) (b) After performing the orientation calibration via the Kabsch algorithm, the data is clearly aligned in both the three-dimensional plot and the time-series plots. (c) Histograms show the distribution of global orientation magnitudes (relative to the reference array orientation) for all individual magnetometers, with the global orientation magnitude of each array labeled and shown by a vertical line.

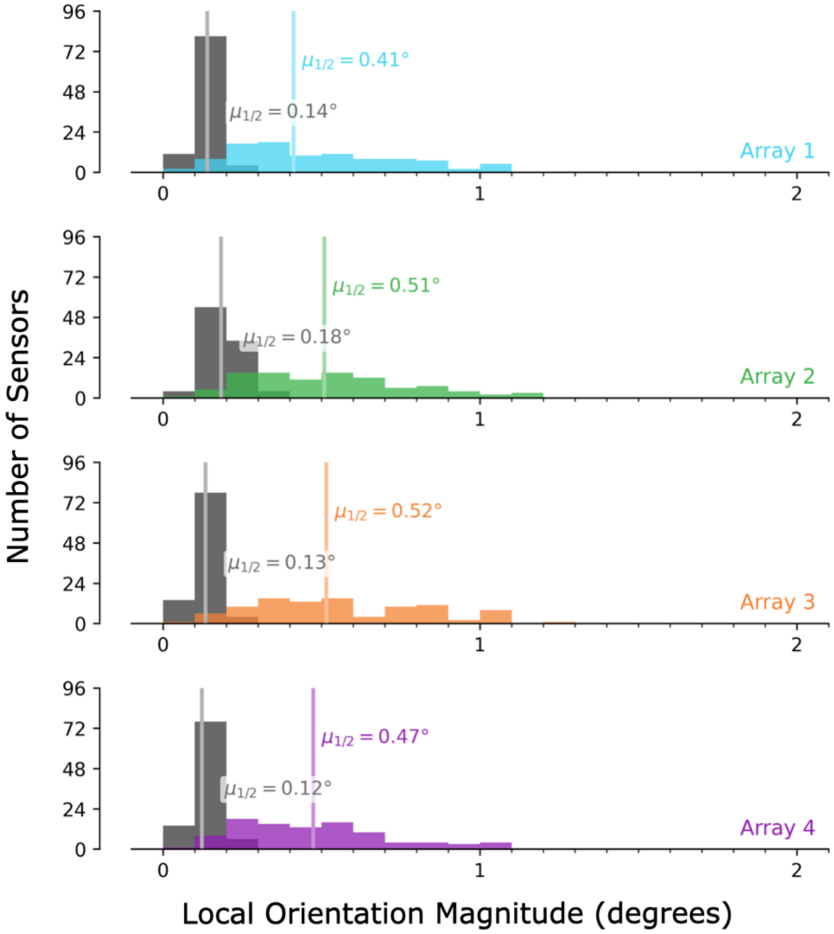


**Figure S7. Repeatability of the magnetometer array orientation calibration**

To test the repeatability of the orientation calibration, we split the full dataset into three parts and performed the orientation calibration on each part separately. We then computed the magnitude of the orientations from these dataset subsets to the orientation from the full dataset to determine an RMS angular error for each magnetometer. The histograms in black show the distribution of RMS angular errors of the individual magnetometers for each array, and are shown next to the distribution of local orientation magnitudes of each array (in respective array colors) for comparison. For ease of comparison, median values are labeled and indicated by vertical lines.

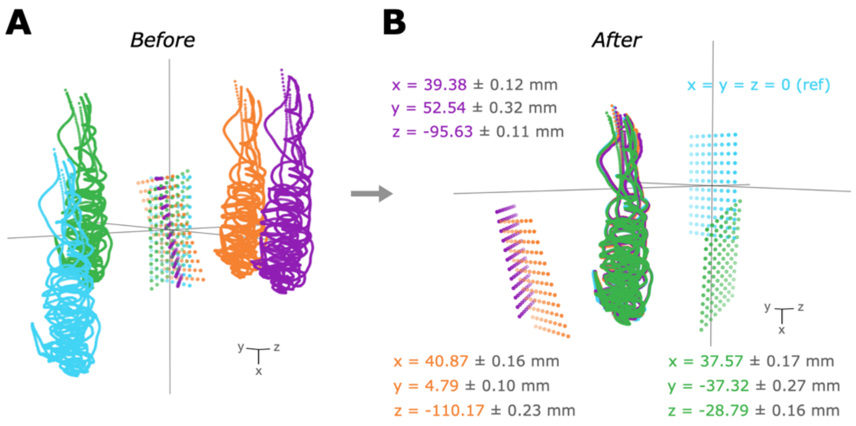


**Figure S8. Translation calibration and repeatability**

The grids of evenly-spaced dots shown are the magnetometer positions, while the curved paths shown give the path of the calibration magnet. (a) To perform position calibration of the magnetometer arrays, we simultaneously independently tracked a calibration magnet from each array in its known orientation relative to the reference array. (b) We then applied a translation-only rigid body transformation to the tracked magnet locations, bringing all magnetometer arrays into a unified global reference frame, by simply calculating the centroids of each point cloud. We then split the full dataset into three parts and performed the translation calibration on each part separately to determine the repeatability of the translation calibration. Translation offsets and standard deviations are shown for each array.

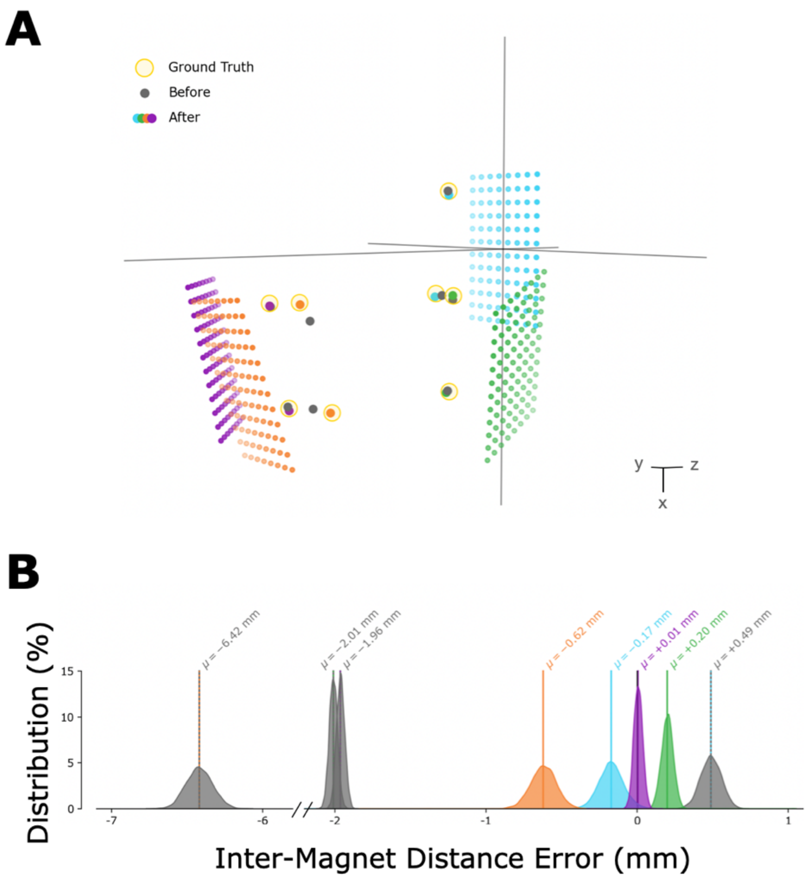


**Figure S9. Static error of tracking before and after adding pose information**

(a) Positions of the magnets as tracked independently from each array with no neighboring magnets present (“Ground Truth"), as tracked independently from each array with all magnets present (“Before"), and as tracked when tracking all magnets from all sensors at once in a unified global frame (“After"). The grids of evenly-spaced dots shown are the magnetometer positions. (b) Distributions of errors in the inter-magnet distance. Errors calculated relative to the ground truth values before and after implementing global tracking.

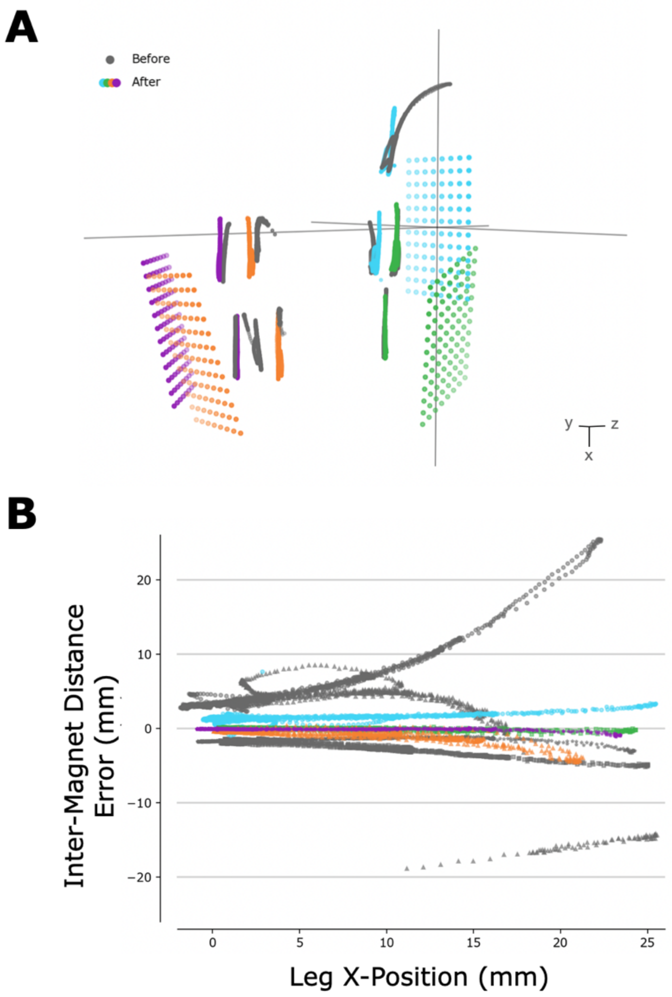


**Figure S10. Inter-magnet distance tracking errors before and after adding pose information**

(a) Positions of the magnets as tracked while the residual limb model was moved vertically relative to the socket, as tracked independently from each array (“Before") and as tracked when tracking all magnets from all sensors at once in a unified global frame (“After"). The grids of evenly-spaced dots shown are the magnetometer positions. (b) Errors in the tracked distance between each magnet pair before versus after implementing global tracking as a function of the vertical position of the residual limb model relative to the socket.

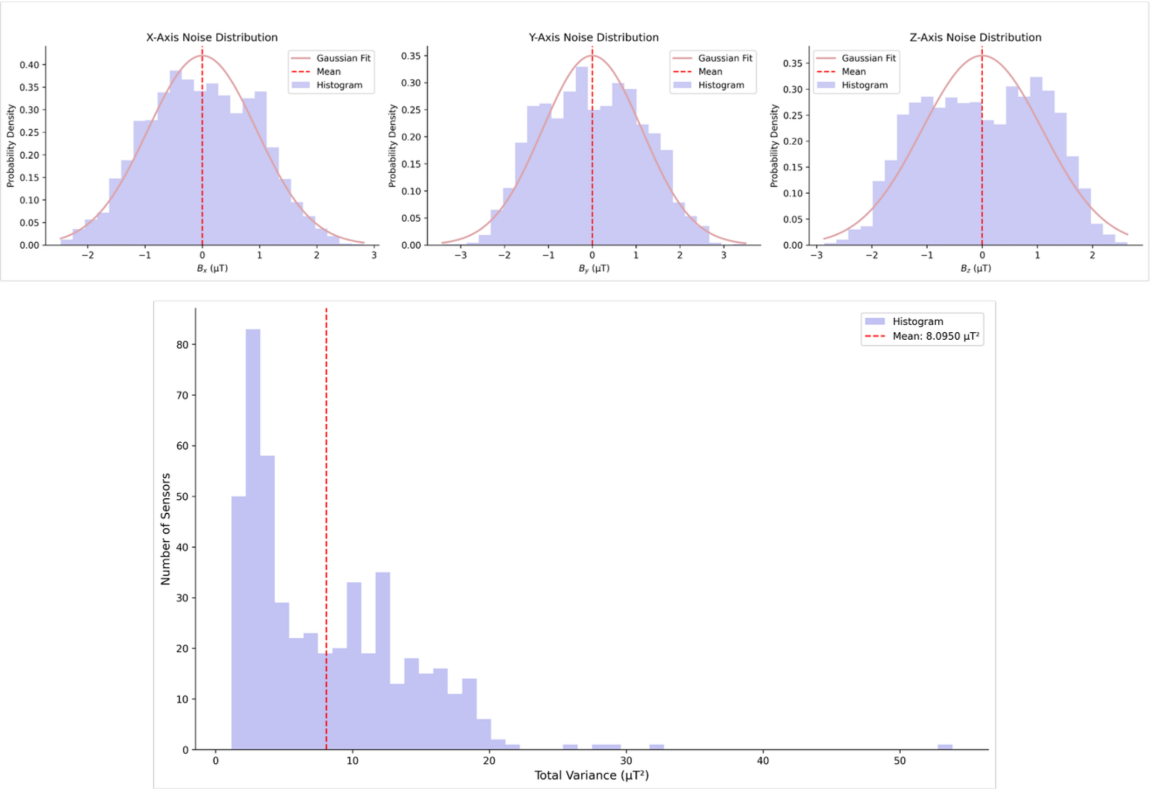


**Figure S11. Magnetic field sensor noise dynamics in steady-state**

The top panels show static noise distributions for X, Y, and Z magnetic field components after hard and soft-iron calibration for a randomly selected sensor, each exhibiting white noise Gaussian characteristics with measurable variance. All three axes demonstrate well-behaved noise profiles with symmetric distributions around their respective means, indicating stable sensor performance. The bottom panel presents the total variance distribution across all sensors, showing a right-skewed distribution with a mean of 8.0950 μT². The majority of sensors exhibit low variance (high precision), while a small subset shows higher variance values, suggesting some sensor-to-sensor variability within the 640-sensor array.

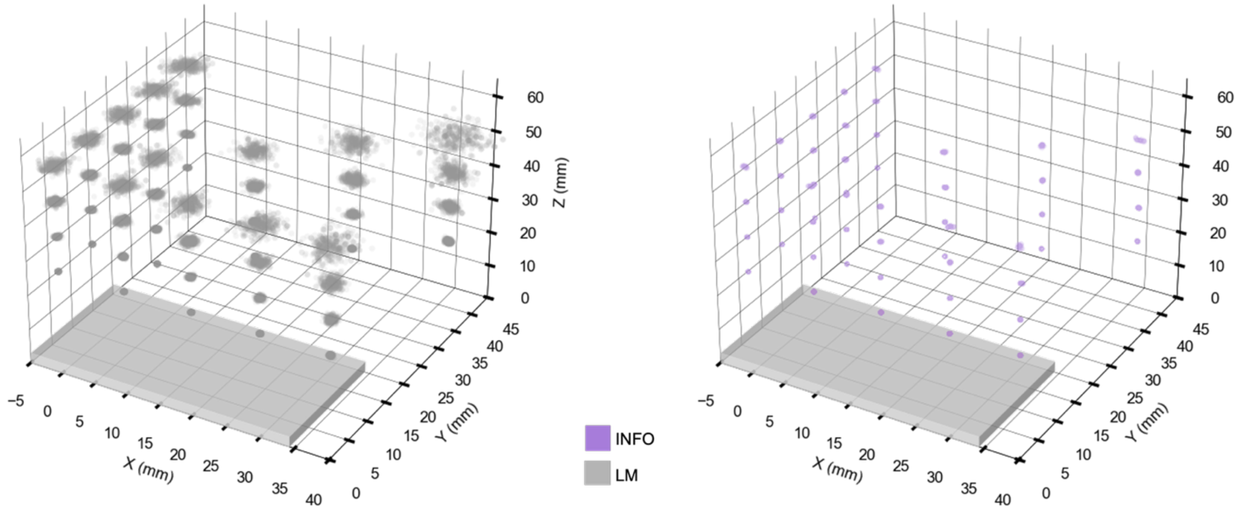


**Figure S12. Single magnet simulation variance**

Scatter plots of position estimation from the single magnet experiment comparing LM (left) and INFO (right) algorithms. The LM method shows dramatically increasing estimation covariance (expanding point-cloud spread) as the magnet distance from the sensor board increases. In contrast, the INFO algorithm maintains consistently low estimation covariance across all magnet distances, demonstrating improved and consistent performance even at extended ranges.

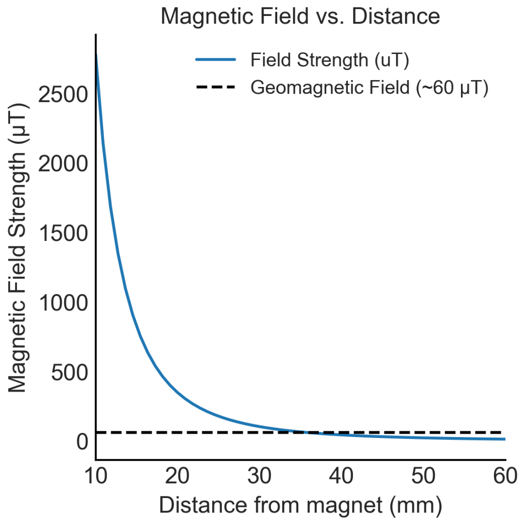


**Figure S13. Magnetic field strength with increasing distance from magnet**

The magnetic field strength B ($\mu T$) exhibits a rapid decay following a $B \propto 1/r^3$ relationship, where r is the distance from the magnetic source. This cubic dependence results in a dramatic reduction in field strength with increasing distance, demonstrating the highly localized nature of the magnetic field. The steep decline highlights the importance of proximity between the sensor and the magnet. At some distance, the field generated from the magnetic point source is lower than the geomagnetic field, creating tracking difficulty if geomagnetic disturbance estimation is poor.

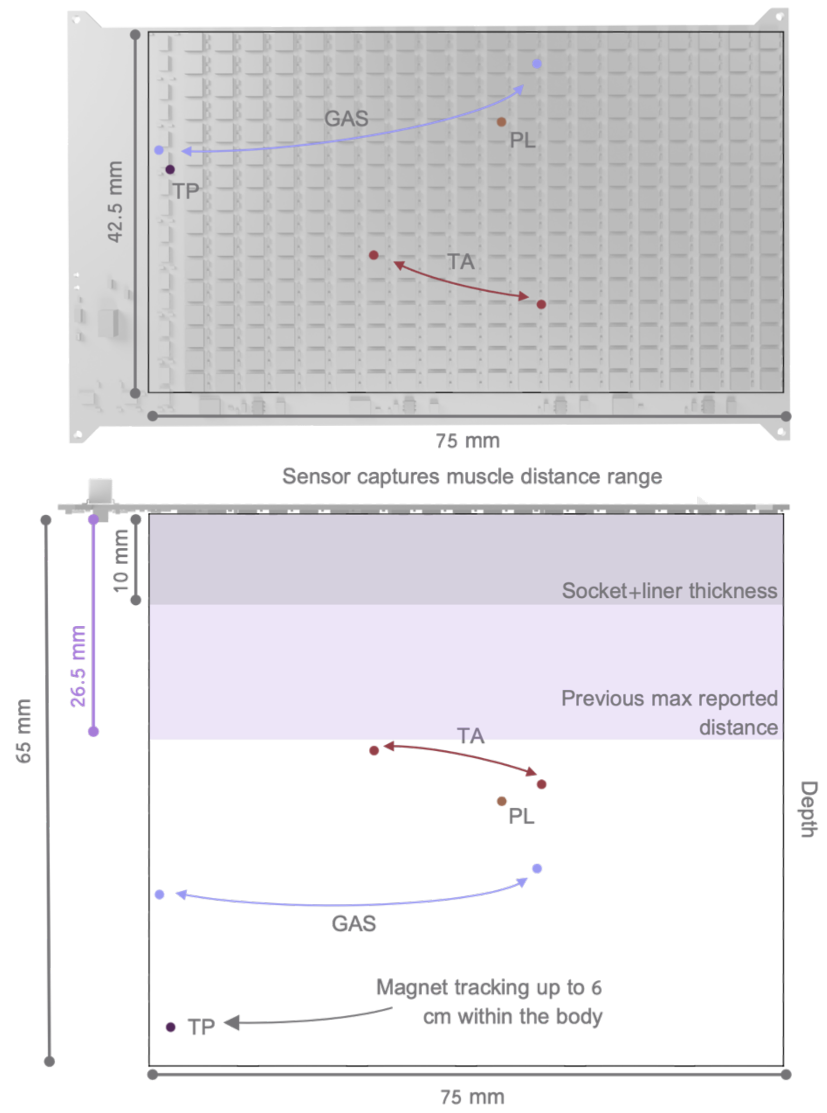


**Figure S14. Tracking depth of magnet within the human body**

The top panel shows a top-down view of the sensor array with tracked static magnet positions for different muscle groups and different sensor boards across Subject 1: gastrocnemius (GAS), tibialis posterior (TP), tibialis anterior (TA), and peroneus longus (PL). The bottom panel presents a side view illustrating tracking depths up to 65 mm within the body, significantly exceeding the previous maximum reported tracking distance of 26.5 mm[2] and socket thickness constraints (indicated by shaded regions). The system successfully captures muscle movement patterns across a range of anatomical locations, with the deepest tracking achieved for the tibialis posterior (TP) at approximately ~60 mm depth.

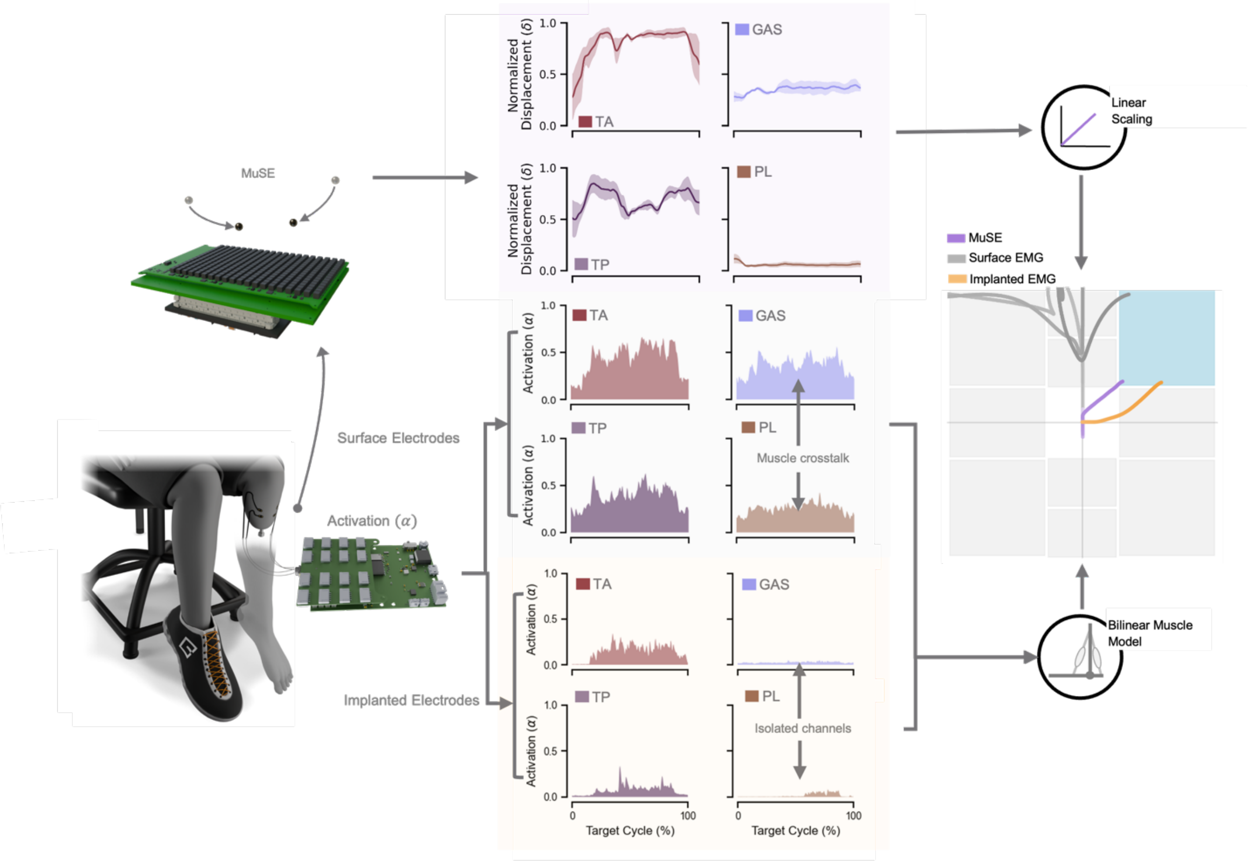


**Figure S15. Muscle cross-talk example in GUI task**

The experimental setup shows recording from the two AMI pairs (TA - tibialis anterior, GAS - gastrocnemius, PL - peroneus longus, TP - tibialis posterior) using three modalities: MuSE, surface EMG, and implanted EMG. Corner target requires co-activation of both the TP and TA muscles. Aggregated data from Subject 1 across three trials of the displayed corner target. Normalized displacement data from MuSE across all muscles is shown during the task. Mean surface EMG in the middle across all muscle channels. Mean implanted EMG at the bottom panel across all muscles. MuSE signals undergoes linear scaling from muscle displacement to ankle position as cursor, while EMG (both surface and implanted) follows a bilinear muscle model to cursor position. Shading indicates SEM for the MuSE measurements.

| **Magnet size (volume mm$^3$)** | **Sensing range** | **Magnet number / Muscle number** | **Magnetometer number** | **Output type** | **Latency** | **Required machine learning compensation** | **Study model** | **Significant improvement over EMG** | **Subject number** | **Ref.** |
|---|---|---|---|---|---|---|---|---|---|---|
| 14.14 mm$^3$ | < 3 cm | 2 / 1 | 96 | Localization (length) | 2.52 ms | - | Turkey | - | - | [2,3] |
| 25.13 mm$^3$ | - | 6 / 3 | 140 | Localization (length) | 23.6 ms | Yes | Human | No | 1 | [4] |
| 261.1 mm$^3$ | - | 3 / 3 | 16 | Raw field | 41.6 ms | Yes | Human | No | 1 | [5] |
| 14.14 mm$^3$ | 6 cm | 6 / 4 | 640 | Localization (length, speed, activation) | 1 ms | No | Human | Yes | 3 | **This work** |

**Table S1. Comparison with other magnetic-based sensing devices.** The muscle state estimator and magnetic beads used in this work are smaller than previous work in humans, provide full muscle state, lower latency and utilize more sensors. In addition, we scaled our previous reported work[2,3] with additional sensors and doubled the tracking depth range.

| **Subject ID** | **Amputation Type** | **Time since amputation (years)** | **Biological sex** | **Height (m)** | **Weight (kg)** | **Time since magnet implantation (years)** |
|---|---|---|---|---|---|---|
| S 1 | AMI | 1.5 | Female | 1.73 | 87.5 | 1.5 |
| S 2 | AMI | 1.3 | Male | 1.7 | 81.1 | 1.3 |
| S 3 | AMI | 2.1 | Male | 1.8 | 78.5 | 0.5 |
| mean ± SEM | | 1.63 ± 0.24 | | 1.74 ± 0.03 | 82.4 ± 2.7 | 1.10 ± 0.31 |

**Table S2. Study population characteristics.** Demographic information for subjects recruited into the clinical study.

**Supplementary Video 1:** Static benchtop testing of different magnet tracking algorithms with MuSE.

**Supplementary Video 2:** Dynamic, real-time magnet tracking with MuSE.

**Supplementary Video 3:** Continuous muscle monitoring through multiple magnets

**Supplementary Video 4:** Real-time control of neuroprosthetic task with MuSE compared to electromyography.
